# Accelerated RNA detection using tandem CRISPR nucleases

**DOI:** 10.1101/2021.03.19.21253328

**Authors:** Tina Y. Liu, Gavin J. Knott, Dylan C. J. Smock, John J. Desmarais, Sungmin Son, Abdul Bhuiya, Shrutee Jakhanwal, Noam Prywes, Shreeya Agrawal, María Díaz de León Derby, Neil A. Switz, Maxim Armstrong, Andrew R. Harris, Emeric J. Charles, Brittney W. Thornton, Parinaz Fozouni, Jeffrey Shu, Stephanie I. Stephens, G. Renuka Kumar, Chunyu Zhao, Amanda Mok, Anthony T. Iavarone, Arturo M. Escajeda, Roger McIntosh, Shin E. Kim, Eli J. Dugan, IGI Testing Consortium, Katherine S. Pollard, Ming X. Tan, Melanie Ott, Daniel A. Fletcher, Liana F. Lareau, Patrick D. Hsu, David F. Savage, Jennifer A. Doudna

## Abstract

Direct, amplification-free detection of RNA has the potential to transform molecular diagnostics by enabling simple on-site analysis of human or environmental samples. CRISPR-Cas nucleases offer programmable RNA-guided recognition of RNA that triggers cleavage and release of a fluorescent reporter molecule^1,2^, but long reaction times hamper sensitivity and speed when applied to point-of-care testing. Here we show that unrelated CRISPR nucleases can be deployed in tandem to provide both direct RNA sensing and rapid signal generation, thus enabling robust detection of ∼30 RNA copies/microliter in 20 minutes. Combining RNA-guided Cas13 and Csm6 with a chemically stabilized activator creates a one-step assay that detected SARS-CoV-2 RNA from nasopharyngeal samples with PCR-derived Ct values up to 29 in microfluidic chips, using a compact imaging system. This Fast Integrated Nuclease Detection In Tandem (FIND-IT) approach enables direct RNA detection in a format amenable to point-of-care infection diagnosis, as well as to a wide range of other diagnostic or research applications.

---

Current strategies for RNA detection in clinical samples based on qRT-PCR (quantitative reverse-transcriptase-polymerase chain reaction) provide high sensitivity (limit of detection of ∼1 molecule/µl) but are too complex to implement for rapid point-of-care testing, an essential component of pandemic control^3–5^. CRISPR-Cas proteins offer a simpler alternative to PCR-based methods due to their programmable, RNA-guided recognition of RNA sequences^6–8^.

Detection takes advantage of the intrinsic enzymatic functions of CRISPR-Cas proteins from Type III and Type VI CRISPR-Cas systems, including the multisubunit effector Csm/Cmr, or the single-protein effector Cas13^1,2,9,10^. Target RNA recognition by the effector triggers multiple-turnover *trans*-cleavage of single-stranded RNA (ssRNA) by either the same protein, in the case of Cas13, or by a separate protein called Csm6, in the case of Csm/Cmr^1,2,11,12^. Enzymatic RNA cleavage generates a fluorescent signal when directed towards a reporter oligonucleotide containing a quencher and dye pair^1,2,13^. Cas13 or Csm/Cmr is capable of reaching an RNA detection sensitivity in the attomolar range in under an hour when coupled to a target sequence amplification procedure, such as reverse transcription-recombinase polymerase amplification (RT-RPA) or reverse transcription-loop-mediated isothermal amplification (RT-LAMP)^1,2,9,10,14,15^. However, addition of RT-RPA or RT-LAMP requires multiple liquid handling steps and/or high-temperature incubation (55-65°C), procedures that are challenging for point-of-care testing^9,10,16–18^. We reported that amplification-free RNA detection using *L. buccalis* Cas13 (LbuCas13) with three guide RNAs could rapidly identify SARS-CoV-2 (severe acute respiratory syndrome coronavirus 2) genomic RNA in patient samples with PCR-derived cycle threshold (Ct) values up to 22, corresponding to ∼1600 copies/µl in the assay^19^. When tested in a mobile phone-based imaging device, this assay detects as low as 200 cp/µl of target RNA within 30 min^19^. However, increased speed and sensitivity of one-pot detection chemistries are still needed to enable their widespread use for point-of-care diagnostics.

Csm6, a dimeric RNA endonuclease from Type III CRISPR-Cas systems, has the potential to boost RNA detection based on its endogenous function in signal amplification^9,10,20^. During CRISPR-Cas interference, activation of Csm or Cmr by viral RNA recognition triggers synthesis of cyclic tetra- or hexaadenylates (cA_4_ or cA_6_) that bind to the Csm6 CRISPR-associated Rossman fold (CARF) and activate its higher eukaryote/prokaryote nucleotide binding (HEPN) domain’s ribonuclease activity^7,11,12,21^. However, diagnostic methods using this cascade were limited to a detection sensitivity of ∼500 fM to 1 nM RNA (∼10^5^-10^9^ cp/µl) without inclusion of RT-LAMP to amplify the target sequence^9,10^. Use of *E. italicus* Csm6 (EiCsm6) in a reaction where Cas13’s target-triggered *trans*-ssRNA cleavage generated linear hexaadenylates with a 2’,3’-cyclic phosphate (A_6_>P, activating ligand of EiCsm6) resulted in low detection sensitivity (1 µM target RNA) and no reduction in detection time^12,20^. Thus, current methods using Csm6 have not resulted in a level of signal amplification that enables CRISPR proteins to directly detect RNA at a concentration relevant for diagnostics.

The limited degree of signal amplification by Csm6 in RNA detection assays could be explained by the observation that Csm6 proteins degrade cA_6_ or cA_4_ over time, causing auto-inactivation^21–25^. Although self-inactivation can be blocked by substituting all the 2’-hydroxyl (2’-OH) groups in the cyclic oligoadenylate with either a 2’-deoxy (2’-H) or 2’-fluoro (2’-F) group, these modifications either completely abolished or led to a 50-fold reduction in Csm6 activation^21,23^. We wondered whether site-selective chemical modification might prevent degradation of the Csm6-activating oligonucleotide while maintaining high-level activation of Csm6. Using the Csm6 protein from the *T. thermophilus* Type III-A system (TtCsm6)^12,26,27^, which recognizes cA_4_ or A_4_>P for activation^12^, we first tested whether 5’-A_3-6_U_6_-3’ oligonucleotides would produce TtCsm6-activating ligands when cleaved by RNA target-bound LbuCas13 (**Fig. 1A, B**). The oligonucleotide A_4_-U_6_ stimulated robust reporter cleavage by TtCsm6 relative to oligonucleotides containing A_3_, A_5_, or A_6_ at the 5’-end **(Fig. 1B)**, consistent with TtCsm6’s preference for a linear activator resembling its natural cA_4_ ligand^11,12^. Furthermore, the reaction requires the LbuCas13-crRNA complex, its target RNA, and the A_4_-U_6_ activator, confirming that TtCsm6 activation is tightly coupled to target RNA detection by LbuCas13 via A_4_-U_6_ activator cleavage (**Fig. 1C**).

**Figure 1.**
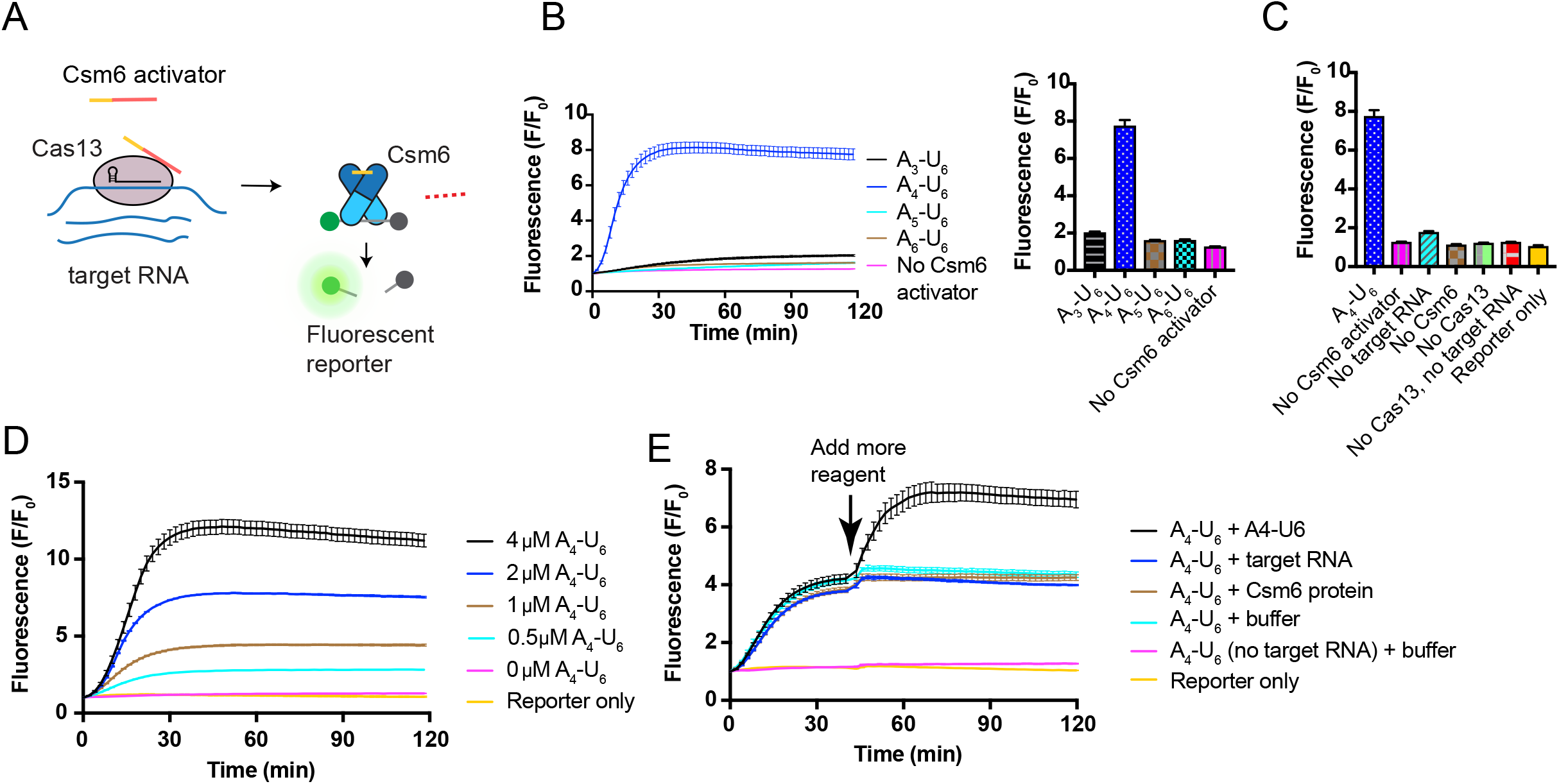
Activation and inactivation of TtCsm6 in an LbuCas13-TtCsm6 assay. **a)** Schematic of TtCsm6 activation by LbuCas13. Binding of an RNA target (blue) by LbuCas13 (pink) leads to activation of *trans*-ssRNA cleavage, which results in trimming of the polyU region (red) from the Csm6 activator. This liberates an oligoadenylate activator with a 2’,3’-cyclic phosphate (yellow) which binds to the Csm6 CARF domains (dark blue) and activates its HEPN (light blue) for cleavage of a fluorophore-quencher RNA reporter. **b)** LbuCas13-TtCsm6 assay with 2 µM Csm6 activator added, containing different lengths of oligoadenylates (A_3_-A_6_ followed by U_6_). LbuCas13 is complexed with a single crRNA sequence, and all reactions contain 200 nM of target RNA, which triggers LbuCas13 to trim off the U’s in the TtCsm6 activator. Normalized fluorescence is plotted (fluorescence value, F, at each timepoint divided by the initial fluorescence, F_0_). The bar graph (right) shows the mean normalized fluorescence and S.E.M. (n = 3) at the assay endpoint of 118 min. **c)** LbuCas13-TtCsm6 assay with controls shown, in which one or more reagents are omitted. Cas13 refers to the Cas13 ribonucleoprotein complex containing both Cas13 protein and its crRNA. Reporter only refers to a reaction with only fluorescent reporter molecules in buffer supplemented with RNase inhibitor. Bar graphs show mean normalized fluorescence and S.E.M. (n = 3) at assay endpoint of 118 min. **d)** As in **b**, but with varying concentrations of the TtCsm6 activator, A_4_-U_6_. **e)** Reactions were initiated as in **b**, but with 1 µM of A_4_-U_6_. The indicated reagents (A_4_-U_6_, Cas13’s target RNA, TtCsm6 protein, or buffer) were then added into the reaction at the time indicated by the arrow. A control reaction is shown without target RNA present in the reaction (no target RNA). Mean normalized fluorescence and S.E.M (n = 3) are plotted over the time course of the reaction.

Activation of TtCsm6 with varying concentrations of A_4_-U_6_ produced an initial burst of fluorescence followed by a plateau within ∼20-30 minutes, with the final fluorescence level proportional to the amount of Csm6 activator present (**Fig. 1D**). Since equal amounts of reporter molecules were present in all reactions, the plateau in the fluorescence signal likely corresponds to a cessation of reporter cleavage by TtCsm6. Addition of A_4_-U_6_ activator to an LbuCas13-TtCsm6 reaction in which the fluorescence had plateaued was shown to rapidly increase reporter cleavage by TtCsm6, while addition of more target RNA or TtCsm6 had no effect relative to a buffer control (**Fig. 1E**). Taken together, these data suggest that Csm6’s A_4_>P ligand is depleted over time, thereby deactivating its RNase activity. Consistent with this conclusion, direct activation of Csm6 with 0.5-2 µM of A_4_>P in the absence of LbuCas13 exhibited a similar pattern of inactivation as that observed for the full LbuCas13-TtCsm6 reaction with A_4_-U_6_ (**Extended Data Fig. 1A**). Liquid chromatography-mass spectrometry (LC-MS) analysis also showed that A_4_>P degrades to A_2_>P following incubation with TtCsm6, suggesting that linear activators, like cyclic oligoadenylates^22–24^, are subject to Csm6-catalyzed cleavage (**Extended Data Table 1**). In the context of the natural cA_4_ ligand, A_4_>P may represent an intermediate on the pathway to full TtCsm6 inactivation (**Fig. 2A**)^22,23^.

**Figure 2.**
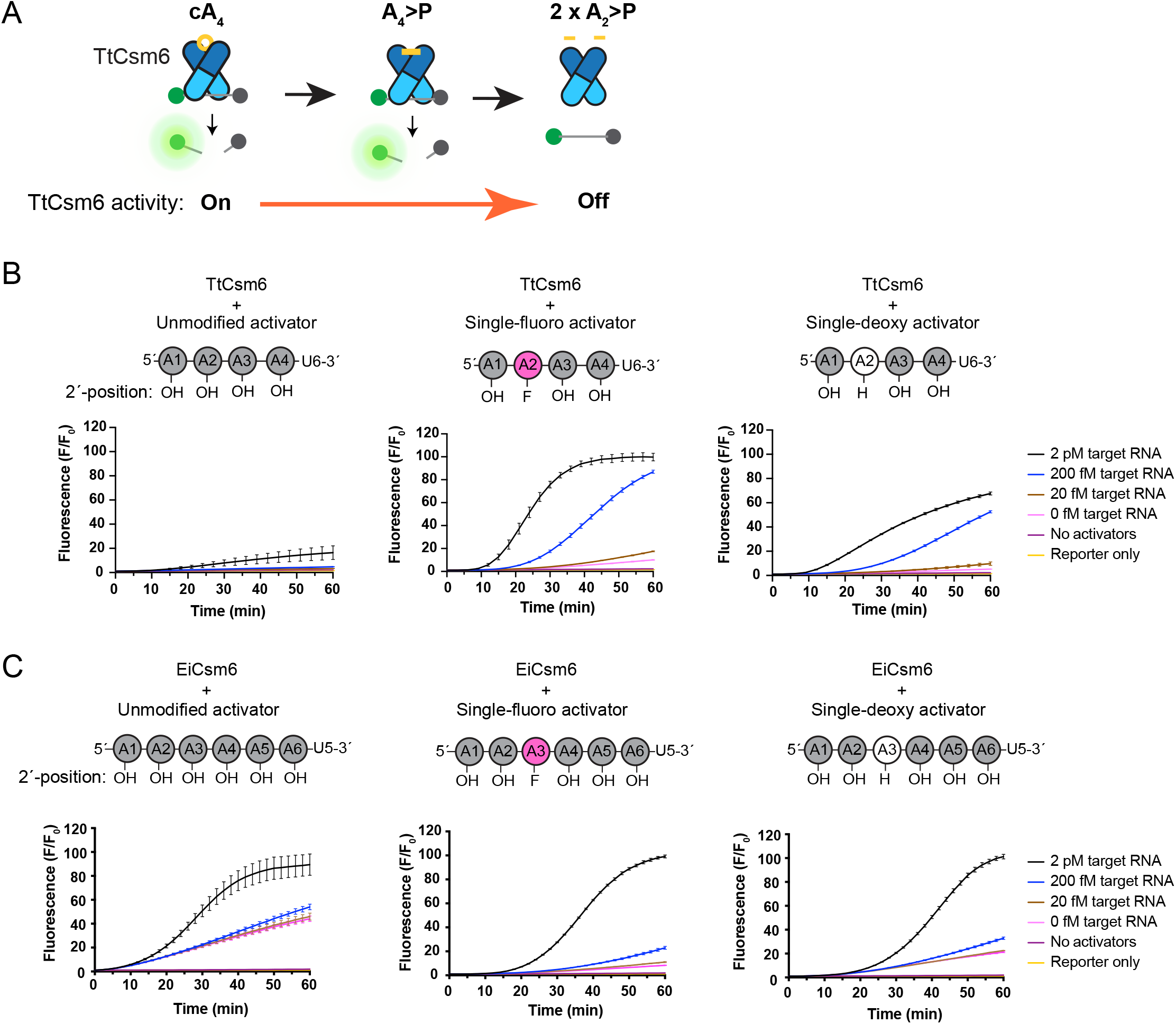
Effect of single 2’-fluoro and 2’-deoxy activator modifications on Csm6-based signal amplification of RNA detection. a) Schematic showing degradation of cA_4_ to A_4_>P and A_2_>P by Csm6’s CARF domain and its effect on RNA cleavage by the HEPN domain. Only binding of the cA_4_ or A_4_>P to the CARF domains (dark blue) of TtCsm6, can activate its HEPN domain (light blue) for ssRNA cleavage. Cleavage of the activator from cA_4_ to A_4_>P and A_2_>P is catalyzed by the CARF domain, and leads to inactivation of the HEPN domain. b) LbuCas13-TtCsm6 reactions with 2 µM of unmodified A_4_-U_6_ (left), or A_4_-U_6_ bearing a single-fluoro modification (center) or single-deoxy modification (right) on the A2 nucleotide. LbuCas13 is loaded with crRNAs 604 and 612, which target an *in vitro* transcribed RNA transcript corresponding to a SARS-CoV-2 genome fragment. Target RNA was added at concentrations from 0-2 pM, and control reactions lacking Csm6 activator and target RNA (“No activators”) or containing only reporter in buffer (Reporter only) are also shown. A schematic showing the position and type of modified nucleotide in the activator is shown above each graph. Adenosines with a 2’-OH, 2’-F, or 2’-H are shown in gray, pink, or white, respectively. Mean normalized fluorescence and S.E.M (n = 3) are plotted over 60 min. c) As in **b**, but with an LbuCas13-EiCsm6 reaction with 0.5 µM of unmodified EiCsm6 activator, A_6_-U_5_ (left), a single-fluoro A_6_-U_5_ (center), and a single-deoxy A_6_-U_5_ (right). The 2’-modification is placed on the A3, rather than A2, to block degradation of A_6_>P to A_3_>P by EiCsm6’s CARF domain^21^.

Mathematical modeling suggested that eliminating activator cleavage could dramatically improve fluorescent signal production in the Cas13-Csm6 detection assay (**Extended Data Fig. 2)**. Mutations in Csm6 or replacement of the 2’-hydroxyl (2’-OH) in cyclic oligoadenylates with 2’-fluoro (2’-F) or 2’-deoxy (2’-H) were previously shown to inhibit activator degradation, but these strategies either drastically lowered or prevented activation of Csm6’s RNA cleavage activity^21,22^. To test whether modified linear activators can increase detection sensitivity and/or speed in a LbuCas13-TtCsm6 assay, we replaced the 2’-hydroxyls of A_4_-U_6_ with either a single or multiple 2’-deoxy, 2’-fluoro, or 2’-O-methyl groups to block cleavage within the A_4_ sequence (**Fig. 2B, Extended Data Fig. 3A-C**). Although inserting multiple modifications into the activators led to slow activation of TtCsm6, substitution of the 2’-hydroxyl of A2 with either a 2’-deoxy or 2’fluoro modification boosted the target RNA detection limit from 2 pM to 20 fM, constituting a 100-fold increase in sensitivity over an unmodified A_4_-U_6_ (**Fig. 2B, Extended Data Fig. 3A-C**). In addition, these singly-modified activators enabled faster distinction between signal and background for reactions measured with 2 pM (detected at ∼10 min), 200 fM (detected at ∼20 min), and 20 fM (detected at ∼40 min) of target RNA (**Fig. 2B**). The single-fluoro A_4_-U_6_ also exhibited improved signal-to-background over the single-deoxy A_4_-U_6_, indicating its superiority in maintaining TtCsm6 activation (**Fig. 2B**).

To confirm that the enhanced signal amplification provided by the singly-modified A_4_U_6_ activators was indeed due to better activation by the A_4_>P ligand, we showed that A_4_>P oligonucleotides containing a 2’-fluoro or 2’-deoxy at the A2 nucleotide could sustain TtCsm6’s activity for longer times compared to the unmodified A_4_>P (**Extended Data Fig. 1A-C**). In addition, incubation of TtCsm6 with the single-fluoro A_4_>P produced A_3_>P, indicating that Csm6-catalyzed degradation of A_4_>P to A_2_>P was blocked by the modification, but that cleavage could occur at other positions (**Extended Data Tables 1, 2**). Interestingly, we found that application of an analogous single-fluoro substitution strategy to EiCsm6 (EiCsm6), an ortholog that uses cA_6_/A_6_>P as an activating ligand^12,21^, did not lead to a significant improvement in the sensitivity or speed of an LbuCas13-EiCsm6 RNA detection assay (**Fig. 2C; Extended Data Fig. 4A, B**). It is possible that the longer A_6_>P activator of EiCsm6 is more susceptible to nucleolytic cleavage relative to the shorter A_4_>P activator of TtCsm6. Taken together, these results demonstrate that the single-fluoro A_4_-U_6_ activator of TtCsm6 improves signal amplification by 100-fold relative to an unmodified activator in a tandem nuclease detection assay with Cas13.

For TtCsm6 and its modified activator to be useful for enhancing RNA detection sensitivity and/or speed, the programmable nature of the CRISPR nuclease used for detection must be preserved. To test this in our Cas13-Csm6 assay, we added TtCsm6 and its single-fluoro activator to an LbuCas13 protein programmed with different crRNA sequences that target the RNA genome of SARS-CoV-2, the causative agent of COVID-19 (**Fig. 3A**). Detection using different crRNA sequences exhibited similar sensitivity and kinetics, showing that this one-step assay is programmable, and thus could potentially be adapted for detection of virtually any RNA sequence. To further improve sensitivity, we complexed LbuCas13 with a pool of eight crRNA sequences, which enables coverage of a wide range of SARS-CoV-2 variants (**Extended Data Table 4**). Using this optimized set of crRNAs, LbuCas13 could detect as few as 63 RNA copies per microliter (cp/µl) of externally validated SARS-CoV-2 RNA in 2 hours (**Fig. 3B**). Inclusion of these eight guides in an LbuCas13-TtCsm6 tandem reaction enabled detection of 31 cp/µl of SARS-CoV-2 RNA in the same time frame (**Fig. 3C**). To determine whether tandem LbuCas13-TtCsm6 could accelerate detection to a time frame better suited to point-of-care testing, we also compared the results of these two detection chemistries after a 20-minute reaction time. While LbuCas13 was unable to detect concentrations ranging from 31-125 cp/µl by 20 minutes, the assay containing both LbuCas13 and TtCsm6 could detect 31 cp/µl within 20 min (p-value < 0.05, **Fig. 3B, C**). Taken together, these results demonstrate that, with optimized crRNAs for LbuCas13 and a chemically stabilized activator for TtCsm6, the tandem CRISPR nuclease assay can detect RNA sequences from an infectious pathogen with both a sensitivity and time to detection that is well-suited for rapid, point-of-care testing.

**Figure 3.**
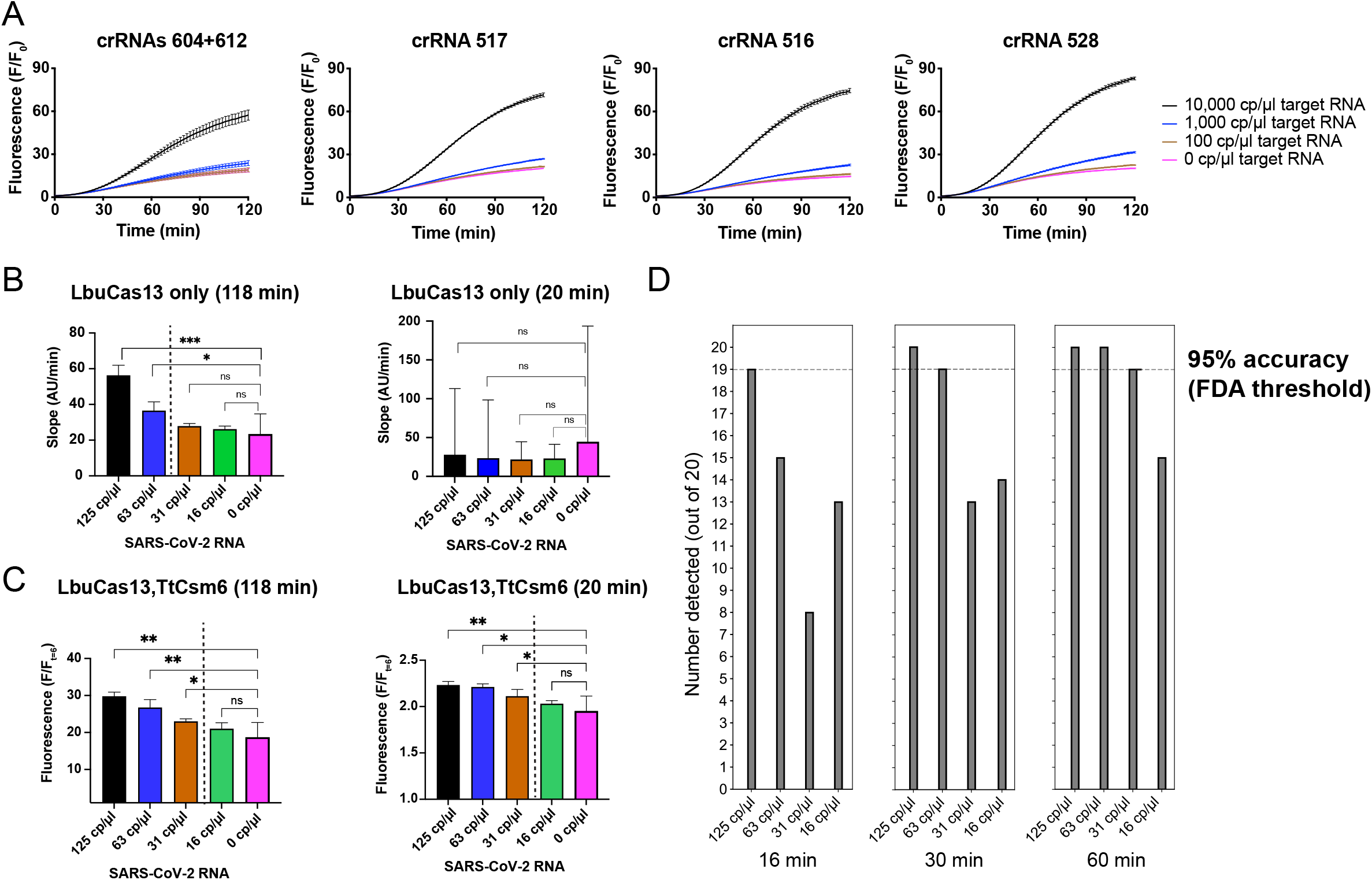
Programmability and benchmarking of the LbuCas13-TtCsm6 assay. a) Testing the programmability of LbuCas13-TtCsm6 with different crRNAs targeting the SARS-CoV-2 genome. In the leftmost panel, LbuCas13 is loaded with two crRNAs, 604 and 612, and in the remaining panels, it is loaded with a single crRNA sequence (516, 517, or 528). Twist synthetic SARS-CoV-2 control RNA was used as the target. Mean normalized fluorescence and S.E.M (n = 3) is plotted over the two-hour time course of the reaction. b) Direct detection of externally validated SARS-CoV-2 RNA control from BEI resources. The slopes of the fluorescent signal generated by reporter cleavage were analyzed at 118 mins and 20 min using LbuCas13 complexed with eight crRNAs targeting the SARS-CoV-2 genome. The mean rate of fluorescence increase, as determined by linear regression, are plotted, with the error bars showing the 95% confidence interval (n = 3). Pairwise comparisons of the slopes to 0 cp/µl by ANCOVA are shown as significant if the p-value < 0.05. Asterisks shown in the figure correspond to the following p-values: * for P ≤ 0.05, ** for P ≤ 0.01, and *** for P ≤ 0.001. Non-significant p-values (P > 0.05) are shown as “ns.” A dotted line indicates the boundary between detected and undetected target RNA concentrations. c) LbuCas13-TtCsm6 detection of BEI SARS-CoV-2 RNA was carried out with eight crRNAs and the single-fluoro TtCsm6 activator. The mean normalized fluorescence (fluorescence at the endpoint divided by fluorescence at 6 min) of three technical replicates was determined at 118 min and 20 min, using t = 6 min as the initial data point for normalization (see **Extended Data Figure 6**). Error bars show the 95% confidence intervals. The significance of pairwise comparisons is represented as in **b** with asterisks to indicate the p-value or “ns” for non-significant p-values. A dotted line indicates the boundary between detected and undetected target RNA concentrations. d) Accuracy of the LbuCas13-TtCsm6 assay assessed over 20 replicates in a plate reader. Individual positive replicates were compared to the 95^th^ percentile of the control distribution to determine if their signal was higher; a difference between the positive replicate and the control distribution was considered significant when p-value < 0.05. The number detected out of 20 replicates is shown at 16, 30, and 60 min for 125 cp/µl, 63 cp/µl, 31 cp/µl and 16 cp/µl of BEI SARS-CoV-2 RNA. The FDA threshold for limit of detection is shown as a dotted line in the bar graphs.

In addition to sensitivity and speed, diagnostic assays must meet the accuracy threshold set by the Food and Drug Administration (FDA) for emergency use authorization, which stipulates that 19 of 20 replicates are detected at ∼1-2 times the limit of detection. To determine if the LbuCas13-TtCsm6 tandem nuclease assay would meet this validation criterion, we ran 20 replicates of our assay around the limit of detection and analyzed individual replicates by comparing them to the 95^th^ percentile of the negative control distribution (**Fig. 3D; see Methods**). Our assay met the 95% accuracy requirement for 20 replicates at 31 cp/µl of viral RNA in 60 min, indicating that this is the limit of detection as defined by the FDA (**Fig. 3D**). In addition, 125 cp/µl and 63 cp/µl of viral RNA were detected with 95% accuracy in as short as 16 min and 30 min, respectively. Taken together, the performance and simplicity of the LbuCas13-TtCsm6 tandem nuclease chemistry creates a Fast Integrated Nuclease Detection In Tandem (FIND-IT) assay that integrates the sensitivity of LbuCas13 with the fast time to detection of the chemically stabilized TtCsm6 activator, while meeting the accuracy required for diagnostic tests.

To demonstrate the feasibility of FIND-IT as a point-of-care test, we developed a detector consisting of a microfluidic chip with reaction chambers, a heating module that maintains the reactions at 37°C, and a compact fluorescence imaging system (**Fig. 4A-C**). The detector, which simultaneously monitors signal from two reaction chambers, is capable of sensing changes in signal as small as 1%, corresponding to a difference of 3 times the Root-Mean-Square-Error (RMSE) in the mean signal. We applied LbuCas13-TtCsm6 reactions containing target RNA (SARS-CoV-2 genomic RNA) or no target RNA to the reaction chambers and monitored the fluorescence signal for one hour (**Fig. 4D**). The signal in a reaction containing 400 cp/µl of extracted SARS-CoV-2 genomic RNA increased non-linearly, resulting in a ∼4.7-fold increase in one hour, while a negative control lacking target RNA exhibited a ∼1.7-fold change (**Fig. 4D, left**). This ∼270% increase in fluorescence of the 400 cp/µl reaction compared to the control contrasts with the ∼3% variation between two negative controls run side-by-side (**Fig. 4D, right**). Comparison of samples to a control reaction run in parallel on the same chip could be used to normalize baseline signal fluctuations between runs. Combined with the substantial difference in output between positive and negative samples, this suggests that the LbuCas13-TtCsm6 reaction is both chemically compatible with the microfluidic chip, and, when implemented in a compact detector, is suitable for use as a point-of-care diagnostic.

**Figure 4.**
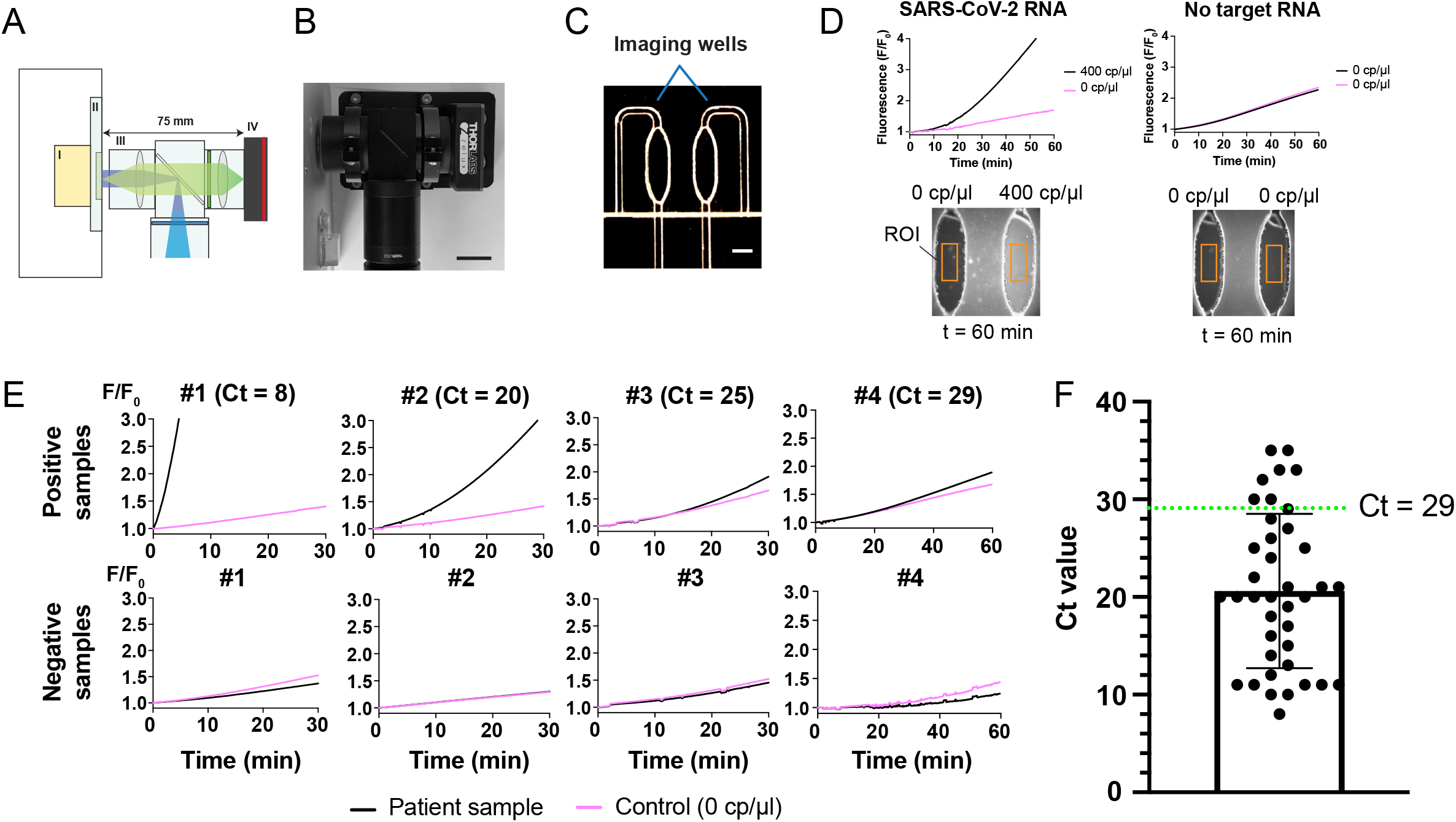
Implementation of FIND-IT on a compact fluorescence detector for testing of COVID-19 patient samples. a) A compact and sensitive fluorescence detector for the LbuCas13-TtCsm6 assay. The heating module (I), sample cartridge (II), imaging optics (III), and camera (IV) are indicated by roman numerals. The size of the imaging optical train is shown above in millimeters (mm). b) Photograph showing the compact fluorescence imaging system. Scale bar is 20 mm. c) Photograph showing the two imaging chambers on the custom microfluidic chip, with a scale bar of 1.75 mm. d) Detection of BEI SARS-CoV-2 genomic RNA on the compact fluorescence detector. Either 400 cp/µl of target RNA or water (0 cp/µl) was added to the reactions, and normalized fluorescence signal is shown over 1 hour. Images below each graph show the final image collected of the imaging chambers. Typical examples of ROIs (regions of interest) selected for integrating fluorescence intensity are shown as orange boxes. e) Detection of SARS-CoV-2 RNA in total RNA extracted from patient samples obtained from the IGI testing laboratory^28^. Positive samples had Ct values ranging from 8 to 29 (top row), and negative patient samples had Ct values >37 (bottom row)^28^. Each sample was run in parallel on the microfluidic chip with a negative control (0 cp/µl), in which water instead of extracted RNA was added to the reaction. Black lines indicate reactions with the patient sample and pink lines indicate the control reactions. Reactions were run for 30-60 min, with images collected every 10 s. f) Scatter dot plot showing the distribution of qRT-PCR Ct values of 39 positive patient samples identified in 296 samples total, obtained from the IGI testing laboratory^28^. A green dotted line indicates the Ct value of 29. Samples with Ct values at and below the green dotted line were detected by the FIND-IT assay in the integrated detector.

To test whether FIND-IT could detect SARS-CoV-2 genomic sequences in total RNA extracted from nasopharyngeal patient samples, we analyzed four positive samples with qRT-PCR-derived Ct values ranging from 8-29, as well as four negative patient samples, on the fluorescence detector (**Fig. 4E, top row)**^28^. FIND-IT detected a signal above the negative control at <5 min for samples with Ct values of 8 and 20, and at ∼20 min for a Ct of 25. For a sample with Ct value of 29, we detected a difference from the control by ∼40 min, consistent with the substantially lower viral RNA concentration corresponding to the higher Ct value **(Extended Data Fig. 5)**. In contrast, all negative patient samples exhibited a signal either equal to or below that of the control over 30-60 minutes of measurements (**Fig. 4E, bottom row**). Based on the Ct values of all COVID-19-positive samples identified in 296 patient samples from the IGI testing laboratory, our assay would detect ∼82% of all positives (**Fig. 4F**)^28^. Combined with upstream RNA extraction, this assay could be deployed as a point-of-care test to capture most individuals at the early, more infectious stages of COVID-19, when Ct values range between 20-30^29^. In addition, based on an estimated conversion for Ct values to copies of RNA, FIND-IT, combined with a sensitive fluorescence detector, can detect approximately ∼6 cp/µl of viral RNA in 40 min (Ct of 29) and 100 cp/µl of viral RNA in 20 min (Ct of 25) (**Extended Data Fig. 5**).

These results establish FIND-IT as a one-step assay that enables rapid RNA detection with sensitivity, accuracy, and adaptability suitable to point-of-care detection of virtually any RNA sequence. FIND-IT demonstrates the value of stabilizing Csm6 nuclease activation and enabling its tandem use with RNA-cleaving CRISPR-Cas proteins for direct and rapid RNA detection^30–33^. This technology could enable practical on-site detection of viral or human RNA in clinical samples, or plant, fungal, or microbial RNA in environmental samples.

## Methods

### Protein purification

The DNA construct encoding *Leptotricia buccalis* Cas13a (LbuCas13a) was codon-optimized for expression in *E. coli*^13^. LbuCas13a expression and purification were carried out as previously described^13^ with modifications, summarized here: N-terminal His_6_-MBP-TEV-tagged or His_6_-SUMO-tagged Cas13a was transformed into *E. coli* Rosetta 2 (DE3) pLysS cells and cultured in terrific broth at 37°C. At an OD_600_ of 0.6–0.8, cultures were cooled on ice for 15 min prior to induction with 0.5 mM isopropyl β-D-1-thiogalactopyranoside (IPTG) and expression overnight at 16°C. Compact cell pellets measuring ∼10 mL in a 50 mL Falcon tube were resuspended in 100 mL of lysis buffer and lysed by sonication. Soluble His_6_-MBP-TEV or His_6_-SUMO Cas13a were clarified by centrifugation at 35,000xg, the supernatant was applied to a 5 mL HiTrap NiNTA column (GE Healthcare) and eluted over a linear imidazole (0.01–0.3 M) gradient via FPLC (ÄKTA Pure). Following overnight dialysis of the peak fractions with TEV or SUMO protease, LbuCas13a was further purified by HiTrap SP (GE Healthcare) ion exchange and, in the case of the MBP-tagged construct, removal of MBP using MBP trap HP columns (GE Healthcare). Finally, size-exclusion chromatography was carried out using a Superdex 200 16/600 column (GE Healthcare) with 1 mM TCEP supplemented into the gel filtration buffer. Peak fractions were pooled, concentrated, and aliquoted into PCR strip tubes before snap freezing in liquid nitrogen.

The codon-optimized sequence encoding amino acid residues 2-467 of TtCsm6 (also known as TTHB152) was cloned into an expression vector that also encodes an N-terminal His_6_-SUMO tag followed by a TEV (Tobacco Etch Virus) cleavage site. The protein was expressed in BL21(DE3) cells, as described previously, and pellets were stored at −20°C^27^. Thawed pellets were resuspended in lysis buffer (20 mM HEPES pH 8.0, 500 mM KCl, 5 mM imidazole, 1 mM TCEP) supplemented with EDTA-free protease inhibitors (Roche) and lysed by sonication. Following clarification of the lysate, the His-SUMO-TtCsm6 protein was immobilized on HIS-Select Nickel Affinity Gel (Sigma) at 4°C, washed with lysis buffer, and eluted using the same buffer supplemented with 250 mM imidazole. TEV protease was added to remove the His-SUMO tag, and the protein was dialyzed overnight at 4°C against buffer containing 25 mM HEPES pH 7.5, 150 mM NaCl, 5% (v/v) glycerol, and 1 mM TCEP in a 10,000 MWCO dialysis cassette. The protein solution was then centrifuged to remove aggregates, concentrated, and subjected to size-exclusion chromatography purification on a Superdex 200 16/600 column (GE Healthcare). Peak fractions were pooled, concentrated to ∼600 µM, and flash-frozen in liquid nitrogen. Working stocks were prepared by diluting the protein to 50 µM in buffer containing 20 mM HEPES (pH 7.5), 200 mM KCl, 5% (v/v) glycerol, 1 mM TCEP and flash-freezing in 5 µl aliquots using liquid nitrogen.

The codon-optimized EiCsm6 gene was cloned into a pET expression vector encoding His_6_-tagged EiCsm6 with an intervening TEV cleavage site (ENLYFQG) by Genscript. The protein was expressed and purified by Shanghai ChemPartner using the following steps. The plasmid encoding His-TEV-EiCsm6 was transformed into BL21(DE3) Star cells and grown in LB broth at 37°C to an OD of ∼0.6. Expression was induced with 0.5 mM IPTG and cultures were transferred to 18°C for 16 h. Cells were resuspended in lysis buffer (20 mM HEPES pH 7.5, 500 mM KCl, 5 mM imidazole) supplemented with EDTA-free protease inhibitors (Roche) and lysed by sonication. The protein was immobilized on a 5-ml Ni^2+^-NTA fast-flow column, washed with lysis buffer, and then eluted with a step gradient of imidazole with fractions at 16%, 50%, and 100% of elution buffer (20 mM HEPES pH 7.5, 500 mM KCl, 500 mM imidazole) collected for further purification. The protein was incubated with TEV protease for 32 h at 25°C to remove the His_6_-tag. Following this, the protein was re-applied to the Ni^2+^-NTA column, washed with 20 mM HEPES pH 7.5, 500 mM KCl and untagged protein was eluted with 20 mM imidazole. Then, the protein was subjected to size-exclusion chromatography in buffer containing 20 mM HEPES pH 7.5, 500 mM KCl, and 5% (v/v) glycerol. Peak fractions were collected and concentrated to 15 mg/ml, and flash-frozen in small aliquots using liquid nitrogen.

### Oligonucleotides

Reporter oligonucleotides, which comprised either a polycytosine sequence for Csm6 assays (C5) or a polyuridine sequence for Cas13 assays (U5) were labeled with a 5’-fluorescein and 3’-Iowa Black moiety. They were ordered HPLC-purified from IDT in bulk (250 nmole to 1 µmole) to avoid batch-to-batch variation in basal fluorescence. For direct comparisons of Csm6 reactions done with different activators or target concentrations, the same batch of reporter was always used. The crRNA guides GJK_073 and R004 were ordered as desalted RNA oligonucleotides from IDT at a scale of 25 nanomoles. Cas13 crRNAs 516, 517, 528, 604, 612, 542, 546, 564, 569, 588, and 596 targeting different regions of the SARS-CoV-2 genome were ordered as desalted oligonucleotides from Synthego at a scale of 5 nanomoles. Unmodified and 2’-modified A_4_>P oligonucleotides were synthesized by IDT at a 250 nmole scale. Sequences of oligonucleotides are listed in **Extended Data Table 3**. The set of eight crRNAs for LbuCas13 (604, 612, 542, 546, 564, 569, 588, and 596) used in Fig. 3B-D are able to detect all published SARS-CoV-2 strains in silico (51631 complete SARS-CoV-2 complete genome sequences were downloaded from NCBI under the taxonomy ID 2697049 as of 03/04/2021). The inclusivity for each guide is listed in **Extended Data Table 4**.

### Preparation of SARS-CoV-2 RNA targets

An *in vitro* transcribed RNA corresponding to a fragment of the SARS-CoV-2 genome was used as a target in **Fig. 2** and **Extended Fig 4**. A gene fragment (gBlock) corresponding to nucleotides 27222-29890 of the SARS-CoV-2 Wuhan-Hu-1 variant genome (MN908947.2) was obtained as a gift from Emily Connelly in the lab of Charles Craik at UCSF. The gBlock was PCR amplified with a 5’ primer bearing an extended T7 promoter sequence (GTCGAAA**TTAATACGACTCACTATAGG**) before separating and extracting the template on an agarose (2% w/v) gel (0.5X TAE buffer) and further purification by phenol-chloroform extraction (pH 8.0) of the purified template and ethanol precipitation. The highly purified and RNase-free template was used in a high yield *in vitro* transcription reaction containing 1X transcription buffer (30 mM Tris-Cl, 25 mM MgCl_2_, 0.01% (v/v) Triton X-100 and 2 mM spermidine, pH 8.1), 5 mM of each NTP, 10 mM DTT, 1 µg/mL pyrophosphatase (Roche), and 100 µg/mL T7 RNA polymerase (purified in house) incubated at 37°C for 4 hours. The reaction was quenched with the addition of 25 units of RNase-free DNase (Promega) at 37°C for 30 mins before the addition of 2 volumes of acidic phenol and subsequent phenol-chloroform extraction. The RNA was then ethanol precipitated and snap-frozen for storage at −80°C.

Twist synthetic SARS-CoV-2 RNA control 2 (Catalog # SKU 102024) was used as a target in **Fig. 3A**. An externally validated genomic SARS-CoV-2 RNA control from BEI resources (NIAID/NIH) was used in **Fig. 3B-D, Fig. 4D**, and **Extended Data Fig. 6** for better comparability to previous studies using this reagent (Lot numbers 70034085 and 70034826). This reagent was deposited by the Centers for Disease Control and Prevention and obtained through BEI Resources, NIAID, NIH: Genomic RNA from SARS-Related Coronavirus 2, Isolate USA-WA1/2020, NR-52285. BEI control RNA was handled in accordance with environmental health and safety regulations for biosafety level 2 materials.

### LbuCas13-TtCsm6 plate reader assays

In **Fig. 1** and **Extended Data Fig. 3**, LbuCas13-TtCsm6 reactions contained 40 nM LbuCas13, 20 nM crRNA (GJK073 or R004), 100 nM TtCsm6, 100-200 pM RNA target (GJK075 or R010), and 200 nM C5 reporter, with either Csm6 activator (A_3-6_U_6_) or DEPC-treated water added for the no-activator control. The reactions were carried out at 37°C in buffer containing 20 mM HEPES (pH 6.8), 50 mM KCl, 5 mM MgCl_2_, 100 µg/ml BSA, 0.01% Igepal CA-630, and 2% glycerol.

For experiments in **Fig. 1E**, the same conditions were used as above except 1 µM of the A_4_-U_6_ activator was added. The reaction was initiated and allowed to proceed until it reached a plateau, at which point 1 µl of either 20 µM A_4_-U_6_, 4 nM target RNA, or 2 µM TtCsm6 protein was added to double the amount of each reagent that was in the reaction. For buffer controls, 1 µl of the reaction buffer was added instead. LbuCas13-crRNA was assembled at a concentration of 1 µM LbuCas13 and 500 nM crRNA for 15 minutes at room temperature. 20-µl plate reader assays were started by mixing 15 µl of a mastermix containing the LbuCas13-crRNA complex and TtCsm6 in buffer with 5 µl of a second mastermix containing RNA target, reporter, and the Csm6 activator in buffer. The protein mastermix was also equilibrated to room temperature for ∼15-20 min prior to starting the reaction. Fluorescence measurements were made every 2 minutes on a Tecan Spark plate reader set with an excitation wavelength of 485 nm, emission wavelength of 535 nm, and z-height optimized to a well containing the reporter molecule in buffer.

In **Figs. 2 and 3**, LbuCas13-TtCsm6 reactions contained 50 nM LbuCas13, 50 nM crRNA, 100 nM TtCsm6, 1 U/µl of Murine RNase inhibitor (New England Biolabs), 2 µM of TtCsm6 activator, and 200 nM C5 reporter, with either target RNA or DEPC water added. The reaction buffer used for these assays contained 20 mM HEPES pH 6.8, 50 mM KCl, 5 mM MgCl_2_, and 5% (v/v) glycerol in DEPC-treated, nuclease-free water (Fisher Scientific or Invitrogen) supplemented with 1 U/µl Murine RNase inhibitor, as previously described^19^. Reactions were started by mixing 15 µl of the protein mastermix, containing LbuCas13, crRNA, TtCsm6, with 5 µl of the activator/reporter mastermix, containing TtCsm6 activator, target RNA, and a fluorescent C5 reporter. Measurements were taken in a plate reader every 2 min at 37°C. In **Figs. 2** and **3A**, measurements were taken on a Biotek plate reader with excitation of 485 nm and emission of 528 nm with z-height of 10.0 mm, and in **Figs. 3C-D**, a Tecan Spark plate reader was used with excitation wavelength of 485 nm and emission wavelength of 535 nm and z-height optimized to a well containing the reporter molecule in buffer.

For LbuCas13-EiCsm6 assays in **Fig. 2** and **Extended Data Fig. 4**, the same buffer and concentrations of LbuCas13, crRNA, and reporter were used as for the LbuCas13-TtCsm6 assays, except 10 nM EiCsm6 and 0.5 µM EiCsm6 activator were added instead of TtCsm6 and its activator. 20 µl reactions were started by addition of a 15-µl LbuCas13/EiCsm6 mastermix to a 5 µl activator/reporter mastermix. Murine RNase inhibitor (NEB) was also included in both mastermixes at 1 U/µl. Measurements were taken every 2 or 3 min at 37°C in a Biotek plate reader, with excitation wavelength of 485 nm, emission wavelength of 52 8nm, a gain of 43, and z-height of 10.0 mm.

All plate reader assays were carried out in low-volume, flat-bottom, black 384-well plates with a non-binding surface treatment (Corning #3820). Fluorescence of Csm6 reactions were normalized where shown by dividing all values by the initial value at t = 0 (F/F_0_), except in **Figs. 3C-D**, where they were normalized to t = 6 min (F/F_6_), to allow for ∼5 min of temperature equilibration to 37°C at the start of the assay (see **Extended Data Fig. 6**). Optically clear ABsolute qPCR Plate Seals (Thermofisher Scientific #AB1170) were used to cover the 384-well plate when doing assays with SARS-CoV-2 genomic RNA. All LbuCas13-Csm6 plate reader experiments in **Figs. 1, 2, 3A, 3C** and **Extended Data Figs. 1, 3, 4** were performed in triplicate. The mean and standard error of the mean (S.E.M.) are plotted in graphs showing the reaction time course.

### Direct activation of TtCsm6 by A_4_>P oligonucleotides

For direct activation experiments, 100 nM TtCsm6 in 1X reaction buffer (20 nM HEPES pH 6.8, 50 mM KCl, 5 mM MgCl_2_, and 5% (v/v) glycerol in DEPC-treated, nuclease-free water) supplemented with 1 U/µl Murine RNase inhibitor was mixed with varying concentrations (0.5-2 µM) of an A_4_>P oligonucleotide. Measurements were taken at 37°C every 2 min in a Tecan Spark plate reader, using an excitation wavelength of 485 nm, emission wavelength of 535 nm, and z-height optimized to a well containing the reporter molecule in buffer. All reactions were performed in triplicate. The mean and standard error of the mean (S.E.M.) are plotted in graphs showing the reaction time course.

### Modeling of the Cas13-Csm6 detection reaction

A kinetic scheme of chemical reactions was created and populated with known kinetic rates and equilibrium constants. Kinetic rates were used where they were known, where only equilibrium rates were known, the forward rates were assumed to be 1 nM^-1^ x sec^-1^ and a reverse rate was chosen to produce the known equilibrium constant. Rates and equilibrium constants were similar to those reported in previous publications^1,12,21,34,35^. For the CARF domain, a K_m_ was known instead of a K_d_, so the CARF k_cat_ was subtracted from the reverse rate to produce an approximate K_d_. The Cas13 background cleavage rate was selected to match the background rate observed in purified LbuCas13 preparations. The full kinetic scheme can be found in the **Supplementary Information (SI)**. This kinetic scheme was then converted into a system of ordinary differential equations modeling the rate of change in the concentration of each reaction component as a function of time and the concentrations of the components using Mathematica. A numerical solution to the system of ordinary differential equations was created with a time step of 0.001 seconds and a total time of 2000 seconds using the Mathematica’s NDSolve at different concentrations of viral target and at different values for the CARF kcat. The full system of ordinary differential equations and starting conditions can be found in **SI**.

### LbuCas13 direct detection experiments and analysis

These assays were performed and analyzed as described previously^19^, with the following modifications. Data were collected on a Tecan Spark plate reader at 37°C with a gain of 87 and a z-height optimized to a well in the plate containing reporter molecule in buffer. Fluorescence was measured every 2 minutes up to 118 min using an excitation wavelength of 485 nm (10 nm bandwidth) and 535 nm emission (20 nm bandwidth). Linear regression and pairwise comparison of slopes using ACNOVA was done using GraphPad Prism 9 as described, considering replicate y-values as individual data points^19^. For determination of the limit of detection at 20 min, we used only data collected in first 20 minutes of the assay. All assays were done in triplicate.

### LC-MS Analysis

The degradation of cA_4_, A_4_>P or fA_4_>P was examined by incubating 5 µM TtCsm6 with 25 µM RNA activator at 37°C for 30 min in a total volume of 15 μl. The reaction was quenched by adding 1 µL 0.5 M EDTA, incubating the sample at 95°C for 5 min and finally centrifuging at 16,000 x g for 5 min to pellet and separate denatured Csm6 protein from its RNA activator. Oligonucleotide samples were then analyzed using a Synapt G2-Si mass spectrometer that was equipped with an electrospray ionization (ESI) source and a BEH C18 ionKey (length: 50 mm, inner diameter: 150 μm, particle size: 1.7 μm, pore size: 130 Å), and connected in line with an Acquity M-class ultra-performance liquid chromatography system (UPLC; Waters, Milford, MA). Acetonitrile, formic acid (Fisher Optima grade, 99.9%), and water purified to a resistivity of 18.2 MΩ·cm (at 25°C) using a Milli-Q Gradient ultrapure water purification system (Millipore, Billerica, MA) were used to prepare mobile phase solvents. Solvent A was 99.9% water/0.1% formic acid and solvent B was 99.9% acetonitrile/0.1% formic acid (volume/volume). The elution program consisted of isocratic flow at 1% B for 2 min, a linear gradient from 1% to 99% B over 2 min, isocratic flow at 99% B for 5 min, a linear gradient from 99% to 1% B over 2 min, and isocratic flow at 1% B for 19 min, at a flow rate of 1.5 μL/min. Mass spectra were acquired in the negative ion mode and continuum format, operating the time-of-flight mass analyzer in resolution mode, with a scan time of 1 s, over the range of mass-to-charge ratio (*m*/*z*) = 100 to 5000. Mass spectrometry data acquisition and processing were performed using MassLynx software (version 4.1, Waters).

### Limit-of-detection analysis for LbuCas13-TtCsm6

For determination of limit of detection using the triplicate data in **Fig. 3C**, a one-tailed t-test with Welch’s correction was used to compare mean fluorescence fold-change of a positive reaction (with initial point being t = 6 min) to that of the control without RNA target, using GraphPad Prism 9. The limit of detection was determined using the triplicate data collected at 20 min and 118 min as timepoints for comparison to LbuCas13 detection assays. Asterisks shown in the figure correspond to the following p-values: * for P ≤ 0.05, ** for P ≤ 0.01, and *** for P ≤ 0.001. The sequences of the crRNAs used are given in **Extended Data Table 3**.

### Limit-of-detection analysis for 20-replicate experiments

For all time-series experiments, data were collected at 2 min intervals and normalized by dividing each replicate by its value at t = 6 min to account for initial signal fluctuations during temperature equilibration (**Extended Data Fig. 6**). Samples were prepared and analyzed in batches of 10 with five experimental samples and five 0 cp/µl negative controls started simultaneously by a multichannel pipette. Analysis was performed at the 16-, 30-, and 60-minute time points. At each of these time points the mean and standard deviation of the 0 cp/µl negative controls within each batch were determined and used to fit a normal distribution. The distribution of negative results was then used to calculate the probability of seeing a point equal to or higher than each experimental sample using the survival function of the normal distribution. This value is equivalent to a one-tailed p-value associated with a null hypothesis in which the experimental value is not greater than the value of the 0 cp/µl negative control. The results are displayed as the fraction of experimental samples out of the total 20 that could be distinguished from the negative control using a. p-value cutoff of 0.05. These cutoffs represent a predicted 5% false positive rate. We did not perform multiple hypothesis correction on this analysis to mirror the analysis that would be performed in a clinical setting, where an individual’s sample would not be corrected based on the number of samples run but where type 1 error (false positive rate) would instead be controlled by setting an appropriate p-value cutoff.

### Experiments using the compact LED-based fluorescence detector

The system camera was focused on and aligned with the imaging wells using an empty cartridge. Then, premixed 60 µl reactions of the LbuCas13-TtCsm6 reaction containing the single-fluoro A_4_-U_6_ activator were loaded directly into inlets leading to the ∼15 µl imaging chambers of a microfluidic chip using a pipette, until the chambers were completely filled. The custom chips were fabricated from poly(methyl methacrylate) and pressure-sensitive adhesive. Reactions were assembled using the same conditions and reagent concentrations as the reactions in **Fig. 3C and 3D**, except a total reaction volume of 60 µl was used. The target RNA volume constituted 10% of the total reaction volume in all reactions testing clinical samples and BEI control SARS-CoV-2 RNA. The reaction chamber temperature was maintained at 37°C with ∼1-2°C variation between runs. Reactions were then imaged every 10 s for 30-60 min using the system camera gain 2 dB and exposure setting of either 150 ms or 100 ms. LED excitation was strobed in synchrony with the camera exposure to avoid photobleaching between data points. ROIs used for signal integration were 150 pixels wide and 400 pixels long, centered in each chamber. For imaging the ∼15 μl sample chambers, we require a fairly large field of view (FOV) and a modest numerical aperture (NA) enabling significant depth of focus without images of adjacent sample wells overlapping. To accomplish this in a relatively low-cost, compact device, we designed a custom system using a pair of eyepieces (Edmund Optics, PN#66-208 and 66-210), yielding a system with NA 0.09, FOV diameter 12.0mm, and magnification (M) of 0.54 (chosen to match the sensor size of the Thorlabs CS165MU1 camera to the FOV; sampling at ≥ Nyquist is unnecessary in this “light-bucket” application). The overall system is compact, with nominal track length (sample to camera) of ∼75mm (**Fig. 4A, B**). Fluorescence filters were from Chroma Technologies, ET470/40x, T495lpxr, and ET535/70m, with excitation provided by a 965mW, 470nm LED (Thorlabs M470L4), providing a maximum of ∼ 225mW into the 12 mm dia. sample FOV in an epi-illumination Kohler geometry. Custom control of the imaging hardware was implemented in MATLAB (2020a), using Thorlabs drivers and SDK (ThorCam) to control the camera acquisition, and serial communication to an Arduino Bluefruit Feather board to electronically trigger the LED illumination.

To accurately compare signals between two samples chambers, we used a prospective correction method to account for nonuniform illumination and background effects^36^. Briefly, the measured pixel intensity of reaction chambers ***I***_*meas*_(*x*) is related to actual signals ***I***_*real*_(*x*) through

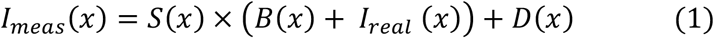

where S is a linear scaling factor that models distortions to an image due to illumination non-uniformity; B is an illumination-dependent background signal that accounts for scattering and background fluorescence from the reaction chambers; and D is an additive zero-light term, which is a result of camera offset and fixed pattern thermal (dark) signal. Prior to actual measurements, we determined S, B, and D by acquiring images of a blank reaction chamber filled with reaction buffer (20 mM HEPES pH 6.8, 50 mM KCl, 5 mM MgCl_2_, and 5% (v/v) glycerol in DEPC-treated, nuclease-free water) and a fluorescent slide, respectively. Experimental images were processed according to equation (1) to retrieve actual signals from each channel.

### Determination of Ct values for patient samples

Human nasopharyngeal swabs were collected, processed, and subjected to RNA extraction and qRT-PCR using primers for SARS-CoV-2 in the IGI testing laboratory, as described previously^28^.

### Determination of Ct values for known concentrations of RNA

Two-fold dilutions of TaqPath COVID-19 Combo Kit positive control (ThermoFisher Scientific) were made in nuclease-free water to generate the indicated copies/μl concentrations. 5 μl of diluted control was added to the multiplexed COVID-19 real-time PCR assay (ThermoFisher Scientific), with a final reaction volume of 12.5μl. Samples were amplified on a QuantStudio6 per the manufacturer’s protocol and analyzed using the Design & Analysis software, v4.2.3 (ThermoFisher Scientific).

### Data and code availability statement

Data that support the findings of this study are available from the corresponding author upon request. Plasmids used in this study will be made available through Addgene. All custom code used for analysis will be made publicly accessible on Github or upon request. Sequences of oligonucleotides used in this study are available in **Extended Data Table 3**.

## Data Availability

Data that support the findings of this study are available from the corresponding author upon request. Plasmids used in this study will be made available through Addgene. All custom code used for analysis will be made publicly accessible on Github or upon request. Sequences of oligonucleotides used in this study are available in Extended Data Table 3.

## Acknowledgments

This work was supported by DARPA under award N66001-20-2-4033. The views, opinions and/or findings expressed are those of the authors and should not be interpreted as representing the official views or policies of the Department of Defense or the U.S. Government. The work was also supported by the Howard Hughes Medical Institute, and the National Institutes of Health (NIH) (R01GM131073 and DP5OD021369 to P.D.H., R01GM127463 to D.F.S., and grant 5R61AI140465-03 to J.A.D., D.A.F., and M.O). A mass spectrometer was also purchased with NIH support (grant 1S10OD020062-01). This work was made possible by a generous gift from an anonymous private donor in support of the ANCeR diagnostics consortium. We also thank the David & Lucile Packard Foundation and the Shurl and Kay Curci Foundation for their generous support of this project. We thank Integrated DNA Technologies and Synthego Corporation for support with oligonucleotide modifications and synthesis, and Shanghai ChemPartner for expression and purification of the EiCsm6 protein. We thank QB3 MacroLabs for subcloning TtCsm6, A. J. Aditham for assistance purifying TtCsm6, and D. Colognori for helpful discussions. J.A.D. is an HHMI investigator. G.J.K. acknowledges support from the NHMRC (Investigator Grant, EL1, 1175568). B.W.T. is supported by a National Science Foundation (NSF) Graduate Fellowship. P.F. was supported by the NIH/NIAID (F30AI143401). M.D.d.L.D. was supported by the UC MEXUS-CONACYT Doctoral Fellowship.

## Author contributions

T.Y.L., J.A.D., and G.J.K. conceived the study. G.J.K, D.C.J.S., and T.Y.L. expressed and purified proteins. T.Y.L. and D.C.J.S. performed all biochemical experiments. T.Y.L., G.J.K., D.C.J.S., E.C., B.W.T., S.J., N.P., and S.A. performed initial experiments optimizing biochemical assays. T.Y.L. designed Csm6 activators with help from G.J.K. and N.P. G.J.K. prepared *in vitro* transcribed gBlock targets. J.J.D. analyzed the 20-replicate experiments and did modeling of the Cas13-Csm6 reaction. S.S., A.B, M.D.d.L.D., N.A.S., M.A., A.R.H., A.M.E., and R.M. developed the compact detector, data normalization method, and software, with M.X.T. and D.A.F. providing supervision. P.F., J.S., S.I.S., C.Z., A.M., G.J.K., and D.C.J.S. identified and analyzed crRNA sequences targeting SARS-CoV-2, with G.R.K., M.O., K.S.P., and L.F.L. providing supervision. A.T.I. performed mass spectrometry data collection and analysis. S.E.K. and E.J.D. provided general experimental support. The IGI Testing Consortium provided clinical samples and associated Ct values. J.R.H. in the IGI Testing Consortium performed experiments converting PCR-derived Ct values to copies of RNA per µl. T.Y.L. wrote the draft of the manuscript with assistance from G.J.K. and J.A.D. All authors edited and approved the manuscript. D.F.S., P.D.H., and J.A.D. obtained funding with significant help from N.P. Overall supervision of the project was provided by T.Y.L, G.J.K., P.D.H., D.F.S., and J.A.D.

## Competing Interests

D.F.S. is a co-founder of Scribe Therapeutics and a scientific advisory board member of Scribe Therapeutics and Mammoth Biosciences. P.D.H. is a cofounder of Spotlight Therapeutics and serves on the board of directors and scientific advisory board, and is a scientific advisory board member to Vial Health and Serotiny. The Regents of the University of California have patents issued and/or pending for CRISPR technologies on which J.A.D., T.Y.L., P.D.H., N.P., D.F.S., D.C.J.S., S.E.K., B.W.T., E.J.C., and G.J.K. are inventors. M.O., P.F., G.R.K., D.F., S.S., and N.A.S. have also filed patent applications related to this work. J.A.D. is a cofounder of Caribou Biosciences, Editas Medicine, Scribe Therapeutics, Intellia Therapeutics and Mammoth Biosciences. J.A.D. is a scientific advisory board member of Caribou Biosciences, Intellia Therapeutics, eFFECTOR Therapeutics, Scribe Therapeutics, Mammoth Biosciences, Synthego, Algen Biotechnologies, Felix Biosciences and Inari. J.A.D. is a Director at Johnson & Johnson and has research projects sponsored by Biogen, Pfizer, AppleTree Partners and Roche.

## Notes

### Author Declarations

Dr. Rebecca Armstrong in the Office for Protection of Human Subjects at UC Berkeley determined that this study did not meet the threshold definition of "human subjects research" set forth in Federal Regulations at 45 CFR 46 or at 21 CFR 50. This determination is based on the understanding that study procedures were limited to secondary data analysis of non-identifiable biospecimens, for which there was no link to identifiable subject information. In addition, the research data in this study will not be held for inspection by nor submitted to the FDA. Accordingly, this project did not fall within the scope of the Committee's responsibilities, and no further approval from the Office is needed at this time.

