## Supplementary Information for "Accelerated RNA detection using tandem CRISPR nucleases"

System of ordinary differential equations and starting conditions for Cas13-Csm6 modeling, related to **Extended Figure 2.**

### 1 Chemical equations for Cas13 to Csm6 transduction model

#### 1.1 Cas13 on-target activity

$$\mathrm{cas}_{13} + \mathrm{guide} \underset{1\,\mathrm{sec}^{-1}}{\overset{1\,\mathrm{nM}^{-1}\mathrm{sec}^{-1}}{\rightleftarrows}} \mathrm{rnp}$$

$$\mathrm{rnp} + \mathrm{target} \underset{1\,\mathrm{sec}^{-1}}{\overset{1\,\mathrm{nM}^{-1}\mathrm{sec}^{-1}}{\rightleftarrows}} \mathrm{rnpActive}$$

$$\mathrm{rnpActive} + \mathrm{a}_4\mathrm{u}_6 \underset{2000\,\mathrm{sec}^{-1}}{\overset{1\,\mathrm{nM}^{-1}\mathrm{sec}^{-1}}{\rightleftarrows}} \mathrm{rnpActivea}_4\mathrm{u}_6$$

$$\mathrm{rnpActivea}_4\mathrm{u}_6 \xrightarrow{200\,\mathrm{sec}^{-1}} \mathrm{rnpActive} + \mathrm{a}_4$$

#### 1.2 Cas13 background activity

$$\mathrm{rnp} + \mathrm{a}_4\mathrm{u}_6 \underset{2000\,\mathrm{sec}^{-1}}{\overset{1\,\mathrm{nM}^{-1}\mathrm{sec}^{-1}}{\rightleftarrows}} \mathrm{rnpa}_4\mathrm{u}_6$$

$$\mathrm{rnpa}_4\mathrm{u}_6 \xrightarrow{4\cdot 10^{-7}\,\mathrm{sec}^{-1}} \mathrm{rnp} + \mathrm{a}_4$$

#### 1.3 Csm6 activity

$$\mathrm{csm}_6 + \mathrm{a}_4 \underset{63\,\mathrm{sec}^{-1} - \mathrm{csm}_6\ \mathrm{carf\ domain\ kcat}}{\overset{1\,\mathrm{nM}^{-1}\mathrm{sec}^{-1}}{\rightleftarrows}} \mathrm{csm}_6\mathrm{Active}$$

$$\mathrm{csm}_6\mathrm{Active} \xrightarrow{\mathrm{csm}_6\ \mathrm{carf\ domain\ kcat}} \mathrm{csm}_6$$

$$\mathrm{csm}_6\mathrm{Active} + \mathrm{reporter} \underset{5000\,\mathrm{sec}^{-1}}{\overset{1\,\mathrm{nM}^{-1}\mathrm{sec}^{-1}}{\rightleftarrows}} \mathrm{csm}_6\mathrm{ActiveReporter}$$

$$\mathrm{csm}_6\mathrm{ActiveReporter} \xrightarrow{0.4\,\mathrm{sec}^{-1}} \mathrm{csm}_6\mathrm{Active} + \mathrm{product}$$

### 2 Cas13 to Csm6 transduction model

#### 2.1 Ordinary differential equations for component concentrations

$$
\begin{aligned}
c'_{\text{a4}}(t) =& -c_{\text{a4}}(t)c_{\text{csm6}}(t) + (63 - \text{csm6 Carf domain kcat})\\
& c_{\text{csm6Active}}(t) + \frac{c_{\text{rnpa4u6}}(t)}{2500000} + 200c_{\text{rnpActivea4u6}}(t),
\end{aligned}
$$

$$
\begin{aligned}
c'_{\text{a4u6}}(t) =& -c_{\text{a4u6}}(t)c_{\text{rnp}}(t) - c_{\text{a4u6}}(t)c_{\text{rnpActive}}(t)+\\
& 2000c_{\text{rnpa4u6}}(t) + 2000c_{\text{rnpActivea4u6}}(t),
\end{aligned}
$$

$$
c'_{\text{cas13}}(t) = c_{\text{rnp}}(t) - c_{\text{cas13}}(t)c_{\text{guide}}(t),
$$

$$
\begin{aligned}
c'_{\text{csm6}}(t) =& -c_{\text{a4}}(t)c_{\text{csm6}}(t)+\\
& (63 - \text{csm6 Carf domain kcat})c_{\text{csm6Active}}(t)+\\
& \text{csm6 Carf domain kcat}c_{\text{csm6Active}}(t),
\end{aligned}
$$

$$
\begin{aligned}
c'_{\text{csm6Active}}(t) =& c_{\text{a4}}(t)c_{\text{csm6}}(t) - (63 - \text{csm6 Carf domain kcat})c_{\text{csm6Active}}(t)-\\
& \text{csm6 Carf domain kcat}c_{\text{csm6Active}}(t) - c_{\text{csm6Active}}(t)\\
& c_{\text{reporter}}(t) + 5000.4c_{\text{csm6ActiveReporter}}(t),
\end{aligned}
$$

$$
c'_{\text{csm6ActiveReporter}}(t) = c_{\text{csm6Active}}(t)c_{\text{reporter}}(t) - 5000.4c_{\text{csm6ActiveReporter}}(t),
$$

$$
c'_{\text{guide}}(t) = c_{\text{rnp}}(t) - c_{\text{cas13}}(t)c_{\text{guide}}(t),
$$

$$
c'_{\text{product}}(t) = 0.4c_{\text{csm6ActiveReporter}}(t),
$$

$$
c'_{\text{reporter}}(t) = 5000c_{\text{csm6ActiveReporter}}(t) - c_{\text{csm6Active}}(t)c_{\text{reporter}}(t),
$$

$$
\begin{aligned}
c'_{\text{rnp}}(t) =& -c_{\text{a4u6}}(t)c_{\text{rnp}}(t) + c_{\text{cas13}}(t)c_{\text{guide}}(t) - c_{\text{rnp}}(t)c_{\text{target}}(t)-\\
& c_{\text{rnp}}(t) + \frac{5000000001c_{\text{rnpa4u6}}(t)}{2500000} + c_{\text{rnpActive}}(t),
\end{aligned}
$$

$$
c'_{\text{rnpa4u6}}(t) = c_{\text{a4u6}}(t)c_{\text{rnp}}(t) - \frac{5000000001c_{\text{rnpa4u6}}(t)}{2500000},
$$

$$
c'_{\text{rnpActive}}(t) = -c_{\text{a4u6}}(t)c_{\text{rnpActive}}(t) + c_{\text{rnp}}(t)c_{\text{target}}(t) - c_{\text{rnpActive}}(t) + 2200c_{\text{rnpActivea4u6}}(t),
$$

$$
c'_{\text{rnpActivea4u6}}(t) = c_{\text{a4u6}}(t)c_{\text{rnpActive}}(t) - 2200c_{\text{rnpActivea4u6}}(t),
$$

$$
c'_{\text{target}}(t) = c_{\text{rnpActive}}(t) - c_{\text{rnp}}(t)c_{\text{target}}(t),
$$

#### 2.2 Starting concentrations of components

$$
\begin{aligned}
c_{\text{a4}}(0) &= 0nM & c_{\text{csm6}}(0) &= 100nM & c_{\text{guide}}(0) &= 13.5078nM\\
c_{\text{a4u6}}(0) &= 2000nM & c_{\text{csm6Active}}(0) &= 0nM & c_{\text{product}}(0) &= 0nM\\
c_{\text{cas13}}(0) &= 13.5078nM & c_{\text{csm6ActiveReporter}}(0) &= 0nM & c_{\text{rnp}}(0) &= 36.4922nM\\
c_{\text{rnpa4u6}}(0) &= 0nM & c_{\text{rnpActive}}(0) &= 0nM & c_{\text{rnpActivea4u6}}(0) &= 0nM\\
c_{\text{target}}(0) &= \text{target}
\end{aligned}
$$
