## Extended Data Tables for "Accelerated RNA detection using tandem CRISPR nucleases"

[10]Biomedical Sciences Graduate Program, University of California, San Francisco, San Francisco, CA 94143, USA
[11]Department of Medicine, University of California, San Francisco, San Francisco, CA 94143, USA
[12]Chan-Zuckerberg Biohub, San Francisco, CA, USA
[13]Center for Computational Biology, University of California, Berkeley, Berkeley, CA, USA;
[14]QB3/Chemistry Mass Spectrometry Facility, University of California, Berkeley, Berkeley, CA, USA
[15]Wainamics Inc., Pleasanton, CA, USA
[16]Department of Epidemiology & Biostatistics, University of California, San Francisco, CA, USA
[17]Howard Hughes Medical Institute, University of California, Berkeley, Berkeley, CA, USA
[18]Department of Chemistry, University of California, Berkeley, Berkeley, CA, USA
[19]Gladstone Institute of Data Science and Biotechnology, Gladstone Institutes, San Francisco, CA, USA
[20]California Institute for Quantitative Biosciences (QB3), University of California, Berkeley, Berkeley, CA, USA.

**EXTENDED DATA TABLES:**

Extended Data Table 1. Analysis of unmodified TtCsm6 activator degradation using LC-MS.

Extended Data Table 2. Analysis of single-fluoro TtCsm6 activator degradation using LC-MS.

Extended Data Table 3. RNA oligonucleotide sequences.

Extended Data Table 4. Inclusivity of crRNAs used for targeting SARS-CoV-2 genome.

**Extended Data Table 1. Analysis of unmodified TtCsm6 activator degradation using LC-MS**

| Sample | $cA_4/A_4$>P (-2) (*m*/*z*=657.15) (Ion counts) | $cA_4/A_4$>P (-1) (*m*/*z*=1315.3) (Ion counts) | $A_2$>P (-1) (*m*/*z*=657.15) (Ion counts) |
|---|---|---|---|
| TtCsm6 + $cA_4$ | ND | ND | 15199 |
| $cA_4$ | 97219 | 26959 | ND |
| TtCsm6 + $A_4$>P | ND | ND | 6432 |
| $A_4$>P | 73529 | 13879 | ND |

ND: Not detected.

$cA_4$: cyclic tetraadenylate

$A_4$>P: linear tetraadenylate with 2´,3´-cyclic phosphate

**Extended Data Table 2. Analysis of single-fluoro TtCsm6 activator degradation using LC-MS**

| Sample | A-fA-AA>P (*m/z* =658.15) (Ion counts) | A-fA-AA>P (*m/z* =1317.3) (Ion counts) | fA-AA>P or A-fA-A>P (*m/z* =988.573) (Ion counts) |
|---|---|---|---|
| TtCsm6 + A-fA-AA>P | ND | ND | 3606 |
| A-fA-AA>P | 14919 | 4875 | 517 |

ND: Not detected.
fA: 2´-fluoro modified adenosine
A>P: adenosine with 2´,3´-cyclic phosphate terminus

### Extended Data Table 3: RNA oligonucleotide sequences

| Name | Sequence | Origin/target (if applicable) | Vendor | Figures used |
|---|---|---|---|---|
| crRNA GJK073 | UAGACCAGCCCAAAAAUGAAGGGCACUAAAACGCAGCGCCUCUUGCAACGAUUAAAUACU | First spacer sequence from the CRISPR array of Lachnospiraceae bacterium genome; targets GJK075 | IDT | Fig. 1 |
| crRNA GJK075 | AGUAUUUAAUCGUUGCAAGAGGCGCUGCUC | Cas13 target RNA (complementary to GJK073) | IDT | Fig. 1 |
| IVT transcript g7 | CAGGUUACUAUAGCAGAGAUAUUACUAAUUAUUAUGAGGACUUUUAAAGUUUCCAUUUGGAAUCUUGAUUACAUCAUAAACCUCAUAAUUAAAAAUUUA<br>UCUAAGUCACUAACUGAGAAUAAAUAUUCUCAAUUAGAUGAAGAGCAACCAAUGGAGAUUGAUUAAACGAACAUGAAAAUUAUUCUUUUCUUGGCACUG<br>AUAACACUCGCUACUUGUGAGCUUUAUCACUACCAAGAGUGUGUUAGAGGUACAACAGUACUUUUAAAAGAACCUUGCUCUUCUGGAACAUACGAGGGC<br>AAUUCACCAUUUCAUCCUCUAGCUGAUAACAAAUUUGCACUGACUUGCUUUAGCACUCAAUUUGCUUUUGCUUGUCCUGACGGCGUAAAACACGUCUAU<br>CAGUUACGUGCCAGAUCAGUUUCACCUAAACUGUUCAUCAGACAAGAGGAAGUUCAAGAACUUUACUCUCCAAUUUUUCUUAUUGUUGCGGCAAUAGUG<br>UUUAUAACACUUUGCUUCACACUCAAAAGAAAGACAGAAUGAUUGAACUUUCAUUAAUUGACUUCUAUUUGUGCUUUUUAGCCUUUCUGCUAUUCCUUG<br>UUUUAAUUAUGCUUAUUAUCUUUUGGUUCUCACUUGAACUGCAAGAUCAUAAUGAAACUUGUCACGCCUAAACGAACAUGAAAUUUCUUGUUUUCUUAG<br>GAAUCAUCACAACUGUAGCUGCAUUUCACCAAGAAUGUAGUUUACAGUCAUGUACUCAACAUCAACCAUAUGUAGUUGAUGACCCGUGUCCUAUUCACU<br>UCUAUUCUAAAUGGUAUAUUAGAGUAGGAGCUAGAAAAUCAGCACCUUUAAUUGAAUUGUGCGUGGAUGAGGCUGGUUCUAAAUCACCCAUUCAGUACA<br>UCGAUAUCGGUAAUUAUACAGUUUCCUGUUUACCUUUUACAAUUAAUUGCCAGGAACCUAAAUUGGGUAGUCUUGUAGUGCGUUGUUCGUUCUAUGAAG<br>ACUUUUUAGAGUAUCAUGACGUUCGUGUUGUUUUAGAUUUCAUCUAAACGAACAAACUAAAAUGUCUGAUAAUGGACCCCAAAAUCAGCGAAAUGCACC<br>CCGCAUUACGUUUGGUGGACCCUCAGAUUCAACUGGCAGUAACCAGAAUGGAGAACGCAGUGGGGCGCGAUCAAAACAACGUCGGCCCCAAGGUUUACC<br>CAAUAAUACUGCGUCUUGGUUCACCGCUCUCACUCAACAUGGCAAGGAAGACCUUAAAUUCCCUCGAGGACAAGGCGUUCCAAUUAACACCAAUAGCAG<br>UCCAGAUGACCAAAUUGGCUACUACCGAAGAGCUACCAGACGAAUUCGUGGUGGUGACGGUAAAAUGAAAGAUCUCAGUCCAAGAUGGUAUUUCUACUA<br>CCUAGGAACUGGGCCAGAAGCUGGACUUCCCUAUGGUGCUAACAAAGACGGCAUCAUAUGGGUUGCAACUGAGGGAGCCUUGAAUACACCAAAAGAUCA<br>CAUUGGCACCCGCAAUCCUGCUAACAAUGCUGCAAUCGUGCUACAACUUCCUCAAGGAACAACAUUGCCAAAAGGCUUCUACGCAGAAGGGAGCAGAGG<br>CGGCAGUCAAGCCUCUUCUCGUUCCUCAUCACGUAGUCGCAACAGUUCAAGAAAUUCAACUCCAGGCAGCAGUAGGGGAACUUCUCCUGCUAGAAUGGC<br>UGGCAAUGGCGGUGAUGCUGCUCUUGCUUUGCUGCUGCUUGACAGAUUGAACCAGCUUGAGAGCAAAAUGUCUGGUAAAGGCCAACAACAACAAGGCCA<br>AACUGUCACUAAGAAAUCUGCUGCUGAGGCUUCUAAGAAGCCUCGGCAAAAACGUACUGCCACUAAAGCAUACAAUGUAACACAAGCUUUCGGCAGACG<br>UGGUCCAGAACAAACCCAAGGAAAUUUUGGGGACCAGGAACUAAUCAGACAAGGAACUGAUUACAAACAUUGGCCGCAAAUUGCACAAUUUGCCCCCAG<br>CGCUUCAGCGUUCUUCGGAAUGUCGCGCAUUGGCAUGGAAGUCACACCUUCGGGAACGUGGUUGACCUACACAGGUGCCAUCAAAUUGGAUGACAAAGA<br>UCCAAAUUUCAAAGAUCAAGUCAUUUUGCUGAAUAAGCAUAUUGACGCAUACAAAACAUUCCCACCAACAGAGCCUAAAAAGGACAAAAAGAAGAAGGC<br>UGAUGAAACUCAAGCCUUACCGCAGAGACAGAAGAAACAGCAAACUGUGACUCUUCUUCCUGCUGCAGAUUUGGAUGAUUUCUCCAAACAAUUGCAACA<br>AUCCAUGAGCAGUGCUGACUCAACUCAGGCCUAAACUCAUGCAGACCACACAAGGCAGAUGGGCUAUAUAAACGUUUUCGCUUUUCCGUUUACGAUAUA<br>UAGUCUACUCUUGUGCAGAAUGAAUUCUCGUAACUACAUAGCACAAGUAGAUGUAGUUAACUUUAAUCUCACAUAGCAAUCUUUAAUCAGUGUGUAACA<br>UUAGGGAGGACUUGAAAGAGCCACCACAUUUUCACCGAGGCCACGCGGAGUACGAUCGAGUGUACAGUGAACAAUGCUAGGGAGAGCUGCCUAUAUGGA<br>AGAGCCCUAAUGUGUAAAAUUAAUUUUAGUAGUGCUAUCCCCAUGUGAUUUUAAUAGCUUCUUAGGAGAAUGACAAAAAAAAAAAAAAAAAA | Fragment of the SARS-CoV2-genome (includes part of the ORF6 gene, and the complete ORF7a, ORF7b, ORF8, N, and ORF10 genes). | N/A | Fig. 2, Extended Data Fig. 4 |
| crRNA 604 | UAGACCACCCCAAAAAUGAAGGGGACUAAAACGGUCCACCAAACGUAAUGCG | Targets N gene in SARS-CoV-2 genome | Synthego Corporation | Fig. 2-4, Extended Data Fig. 4, 6 |
| crRNA 612 | UAGACCACCCCAAAAAUGAAGGGGACUAAAACUUUGCGGCCAAUGUUUGUAA | Targets N gene in SARS-CoV-2 genome | Synthego Corporation | Fig. 2-4, Extended Data Fig. 4, 6 |
| crRNA 542 | UAGACCACCCCAAAAAUGAAGGGGACUAAAACAAACUACGUCAUCAAGCCAA | Targets Orf1a gene in SARS-CoV-2 genome | Synthego Corporation | Fig. 3B-C, Fig. 4, Extended Data Fig. 6 |
| crRNA 546 | UAGACCACCCCAAAAAUGAAGGGGACUAAAACCACAGUCAUAAUCUAUGUUA | Targets Orf1a gene in SARS-CoV-2 genome | Synthego Corporation | Fig. 3B-C, Fig. 4, Extended Data Fig. 6 |
| crRNA 564 | UAGACCACCCCAAAAAUGAAGGGGACUAAAACUCACACUUUUCUAAUAGCAU | Targets Orf1ab gene in SARS-CoV-2 genome | Synthego Corporation | Fig. 3B-C, Fig. 4, Extended Data Fig. 6 |
| crRNA 569 | UAGACCACCCCAAAAAUGAAGGGGACUAAAACUGUAAGAUUAACACACUGAC | Targets S gene in SARS-CoV-2 genome | Synthego Corporation | Fig. 3B-C, Fig. 4, Extended Data Fig. 6 |
| crRNA 588 | UAGACCACCCCAAAAAUGAAGGGGACUAAAACUUAAUUGUGUACAAAAACUG | Targets S gene in SARS-CoV-2 genome | Synthego Corporation | Fig. 3B-C, Fig. 4, Extended Data Fig. 6 |
| crRNA 596 | UAGACCACCCCAAAAAUGAAGGGGACUAAAACCAGUUGUGAUGAUUCCUAAG | Targets Orf8 gene in SARS-CoV-2 genome | Synthego Corporation | Fig. 3B-C, Fig. 4, Extended Data Fig. 6 |
| crRNA 516 | UAGACCACCCCAAAAAUGAAGGGGACUAAAACCAAAAGCCUCAACACGUAGA | Targets Orf1a gene in SARS-CoV-2 genome | Synthego Corporation | Fig. 3A |
| crRNA 517 | UAGACCACCCCAAAAAUGAAGGGGACUAAAACCAUGUUGAAACAAGUAACUC | Targets Orf1a gene in SARS-CoV-2 genome | Synthego Corporation | Fig. 3A |
| crRNA 528 | UAGACCACCCCAAAAAUGAAGGGGACUAAAACUCAACGAUGUAAGAAGACUG | Targets Orf1a gene in SARS-CoV-2 genome | Synthego Corporation | Fig. 3A |
| crRNA R004 | AUUUAGACCACCCCAAAAAUGAAGGGGACUAAAACACAGAUAUAGCCUGGUGGUUCAGGC | Targets R004 | IDT | Extended Data Fig. 3 |
| crRNA R010 | AAAAGCCUGAACCACCAGGCUAUAUCUG | Cas13 target RNA (complementary to R004) | IDT | Extended Data Fig. 3 |
| C5 reporter | 6-FAM-CCCCC-Iowa Black® | Fluorescent reporter RNA for Csm6 | IDT | Figs. 1-4, Extended Data Figs. 1, 3, 4, 6 |
| U5 reporter | 6-FAM-UUUUU-Iowa Black® | Fluorescent reporter RNA for Cas13 | IDT | Fig. 3B |
| TtCsm6 activator 497 | A-fA-AAUUUUUU | TtCsm6 single-fluoro activator | IDT | Fig. 2-4, Extended Data Fig. 6 |
| TtCsm6 activator 470 | A-dA-AAUUUUUU | TtCsm6 single-deoxy activator | IDT | Fig. 2 |
| TtCsm6 activator 142 | AAAAUUUUUU | TtCsm6 unmodified activator | IDT | Fig. 1,2 |
| EiCsm6 Activator 767 | AA-fA-AAAUUUUU | EiCsm6 single-fluoro activator | IDT | Fig. 2, Extended Data Fig. 4 |
| EiCsm6 Activator 766 | AA-dA-AAAUUUUU | EiCsm6 single-deoxy activator | IDT | Fig. 2, Extended Data Fig. 4 |
| EiCsm6 Activator 768 | AAAAAAUUUUU | EiCsm6 unmodified activator | IDT | Fig. 2, Extended Data Fig. 4 |
| TtCsm6 activator 837 | AAAA>P | TtCsm6 unmodified activator | IDT | Extended Data Fig. 1A |
| TtCsm6 activator 839 | A-fA-AA>P | TtCsm6 single-fluoro activator | IDT | Extended Data Fig. 1B |
| TtCsm6 activator 838 | A-dA-AA>P | TtCsm6 single-deoxy activator | IDT | Extended Data Fig. 1C |
| TtCsm6 activator 374 | fA-fA-fAAUUUUUU | TtCsm6 triple-fluoro activator | IDT | Extended Data Fig. 3A |
| TtCsm6 activator 373 | dA-dA-dAAUUUUUU | TtCsm6 triple-deoxy activator | IDT | Extended Data Fig. 3B |
| TtCsm6 activator 372 | mA-mA-mAAUUUUUU | TtCsm6 triple-O-methyl activator | IDT | Extended Data Fig. 3C |
| TtCsm6 cyclic activator | cA4 | TtCsm6 cyclic oligoadenylate activator | Axxora, LLC | Extended Data Tables 1, 2 |

**Guide region is underlined in crRNA sequences**
**6-FAM: 6-carboxyfluorescein**
**Iowa Black: FAM quencher moiety (IDT)**
**fA: 2´-fluoro adenosine**
**dA: 2´-deoxy adenosine**
**mA: 2´-O-methyl adenosine**
**A>P: adenosine with 2´,3´-cyclic phosphate**
**cA4: cyclic tetraadenylate**

#### Extended Data Table 4. Inclusivity of crRNAs used for targeting SARS-CoV-2 genome

| Sequence name | Inclusivity | crRNA sequence | Target Sequence |
|---|---|---|---|
| crRNA_542 | 51485 | AAACUACGUCAUCAAGCCAA | TTGGCTTGATGACGTAGTTT |
| crRNA_546 | 51383 | CACAGUCAUAAUCUAUGUUA | TAACATAGATTATGACTGTG |
| crRNA_564 | 51527 | UCACACUUUUCUAAUAGCAU | ATGCTATTAGAAAAGTGTGA |
| crRNA_569 | 50313 | UGUAAGAUUAACACACUGAC | GTCAGTGTGTTAATCTTACA |
| crRNA_588 | 51602 | UUAAUUGUGUACAAAAACUG | CAGTTTTTGTACACAATTAA |
| crRNA_596 | 50705 | CAGUUGUGAUGAUUCCUAAG | CTTAGGAATCATCACAACTG |
| crRNA_604 | 51402 | GGUCCACCAAACGUAAUGCG | CGCATTACGTTTGGTGGACC |
| crRNA_612 | 51372 | UUUGCGGCCAAUGUUUGUAA | TTACAAACATTGGCCGCAAA |
