## Extended Data Figures for "Accelerated RNA detection using tandem CRISPR nucleases"

##### Figure Legends:

**Extended Data Figure 1: Direct activation of TtCsm6 by $A_4$>P oligonucleotides.**

a) Activation of TtCsm6 by $A_4$>P added at varying concentrations. Mean raw fluorescence intensity with error bars indicating S.E.M. (n = 3) is shown in arbitrary units (AU). The schematic on the right is a representation of the $A_4$>P, with adenosines numbered A1-A4 from 5′ to 3′.

b) As in **a**, but showing activation of TtCsm6 by single fluoro-modified $A_4$>P at varying concentrations. The schematic on the right is as in **a**, but with the modified nucleotide colored pink.

c) As in **a**, but showing activation of TtCsm6 by single deoxy-modified $A_4$>P. The schematic on the right is as in **a**, but with the modified nucleotide colored white.

**Extended Data Figure 2: Modeling of the Cas13-Csm6 detection reaction in the presence and absence of activator degradation**

a) A graph showing the modeled kinetics of reporter cleavage by the HEPN domain of Csm6 when 0-1 pM target RNA is present and the CARF domain is able to cleave the $A_4$>P oligonucleotide generated by Cas13 cleavage of the $A_4$-$U_6$ activator (CARF $k_{cat}$ = 0.05).

b) As in **a**, but in a situation where the CARF domain is unable to cleave the $A_4$>P oligonucleotide generated by Cas13 cleavage of the $A_4$-$U_6$ activator (CARF $k_{cat}$ = 0).

**Extended Data Figure 3: Triple-modified TtCsm6 activators lead to slow kinetics of reporter cleavage in a LbuCas13-TtCsm6 detection assay.**

**a)** LbuCas13-TtCsm6 reaction with crRNA R004, 100 pM of a complementary target RNA (R010) and 4 µM of the triple-fluoro $A_4$-$U_6$ activator. Mean normalized fluorescence and S.E.M (n = 3) are plotted over 120 min. Controls without target RNA, TtCsm6 activator, and TtCsm6 protein are shown, as well as a reaction showing the reporter in buffer (Reporter only). Positions of the 2´-fluoro modifications in the activator are shown in the schematic (right).

**b)** As in **a** but with 4 µM triple-deoxy $A_4$-$U_6$ to activate TtCsm6. Positions of the 2´-deoxy modifications in the activator are shown in the schematic (right).

**c)** As in **a** but with 4 µM triple-O-methyl $A_4$-$U_6$ to activate TtCsm6. Positions of the 2´-O-methyl (O-Me) modifications in the activator are shown in the schematic (right).

**Extended Data Figure 4: LbuCas13-EiCsm6 detection is not improved by single 2´-fluoro modified activators at high target RNA concentrations.**

a) Detection assay using LbuCas13 complexed with crRNAs 604 and 612, and EiCsm6 with 0.5 µM of a $A_6$-$U_5$ activator[20]. An *in vitro* transcribed RNA corresponding to a fragment of the SARS-CoV-2 genome was added at concentrations ranging from 1.6 pM to 160 pM. Mean normalized fluorescence and S.E.M (n = 3) are plotted over 120 min. A schematic of the activator and the position of the modification is shown above the graph.

b) As in **a** but with a single-fluoro $A_6$-$U_5$ activator. The position of the modification in the activator is shown in the schematic above the graph.

**Extended Data Figure 5: Standard curve of Ct values versus concentration**

a) Ct values obtained from qRT-PCR for a dilution series of the positive control RNA provided by the Thermo TaqPath combo kit. The highest concentration of sample added to the reaction is 1000 cp/µl and other sample RNA concentrations were obtained by diluting the concentrated sample two-fold down to ~2 cp/µl. Primers for the N gene of SARS-CoV-2 were used for amplification. Mean Ct value and S.D. (n = 2) are graphed for each concentration of RNA tested.

b) As in **a** but with primers for the Orf1ab gene of SARS-CoV-2.

c) As in **a** but with primers for the S gene of SARS-CoV-2.

**Extended Data Figure 6: Effect of normalizing fluorescence relative to different timepoints in a plate reader assay.**

Graphs of 10 individual replicates from the LbuCas13-TtCsm6 assay in **Fig. 3D** using eight crRNAs targeting the SARS-CoV-2 genome, the single-fluoro TtCsm6 activator, and 125 cp/µl of BEI SARS-CoV-2 RNA as the target. Black curves indicate replicates with target RNA present, and pink curves indicate control replicates without target RNA. Comparison of the normalization at 0, 4, and 6 min (t = 0, 4, 6) shows that normalizing a few minutes after the start of the plate reader assay reduces the effect of initial signal fluctuations on the normalization.

### Extended Data Figure 1

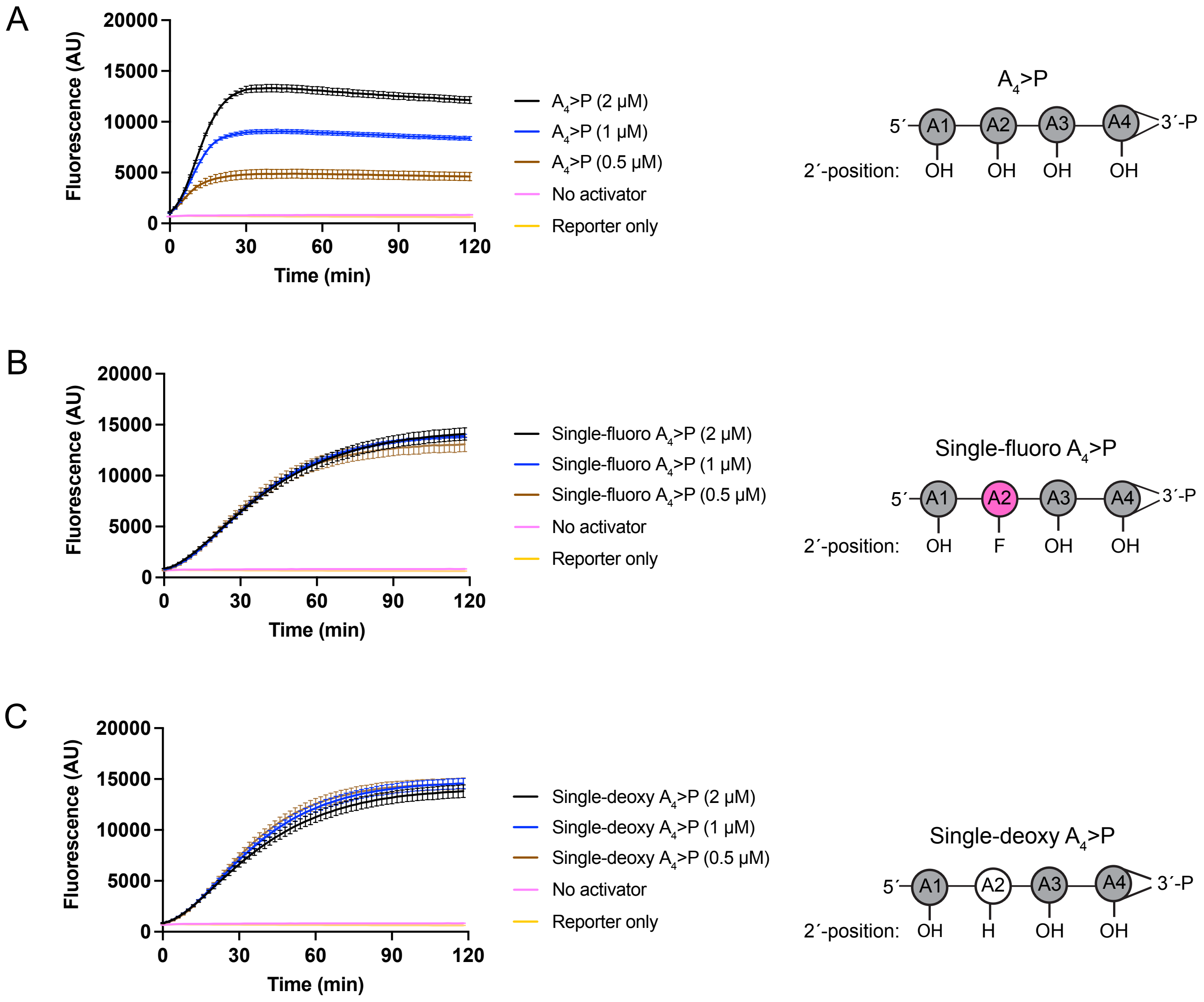

Extended Data Figure 2

A

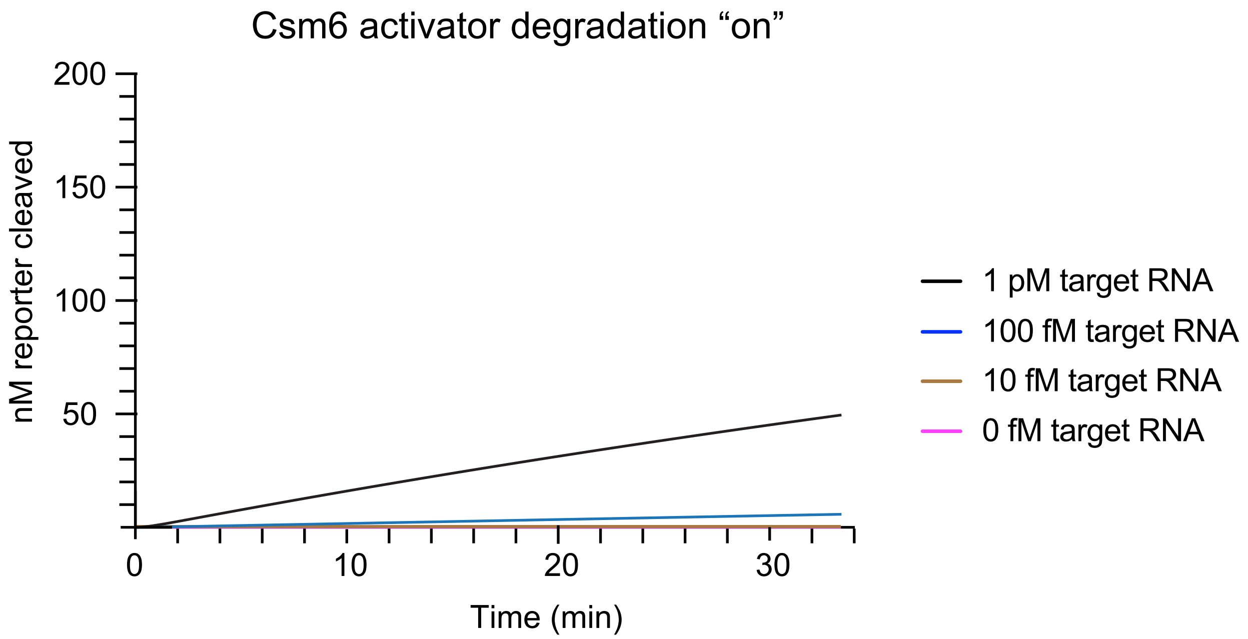


B

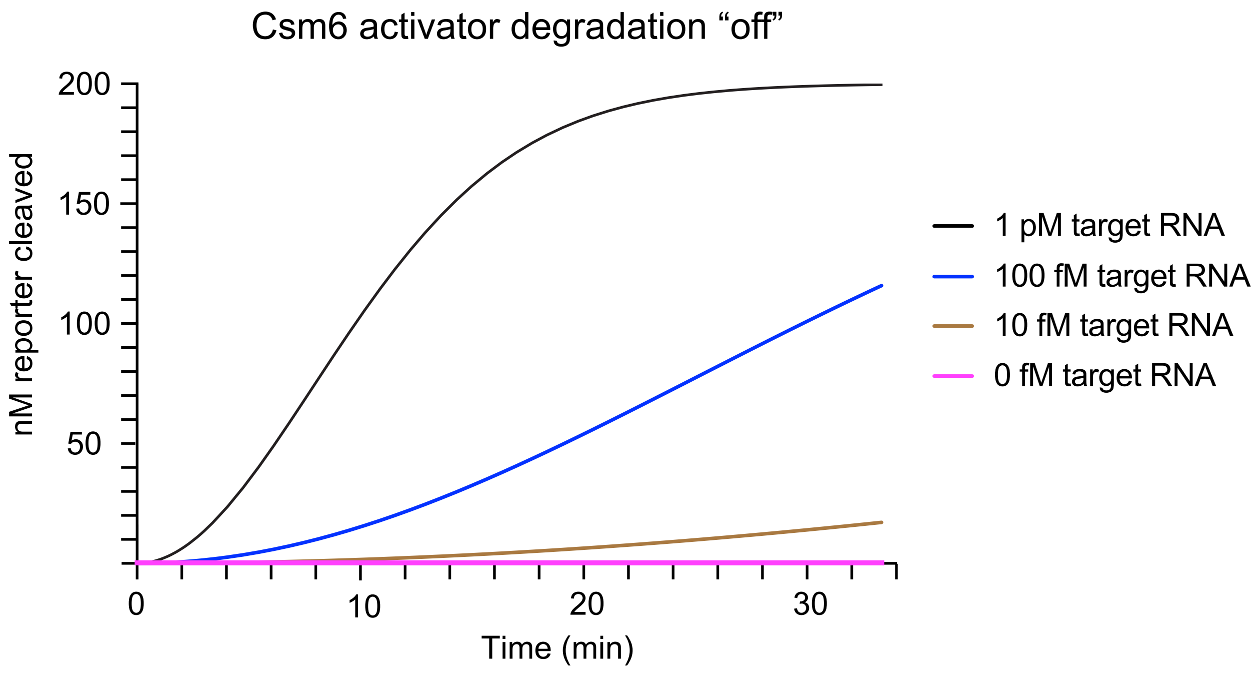

Extended Data Figure 3

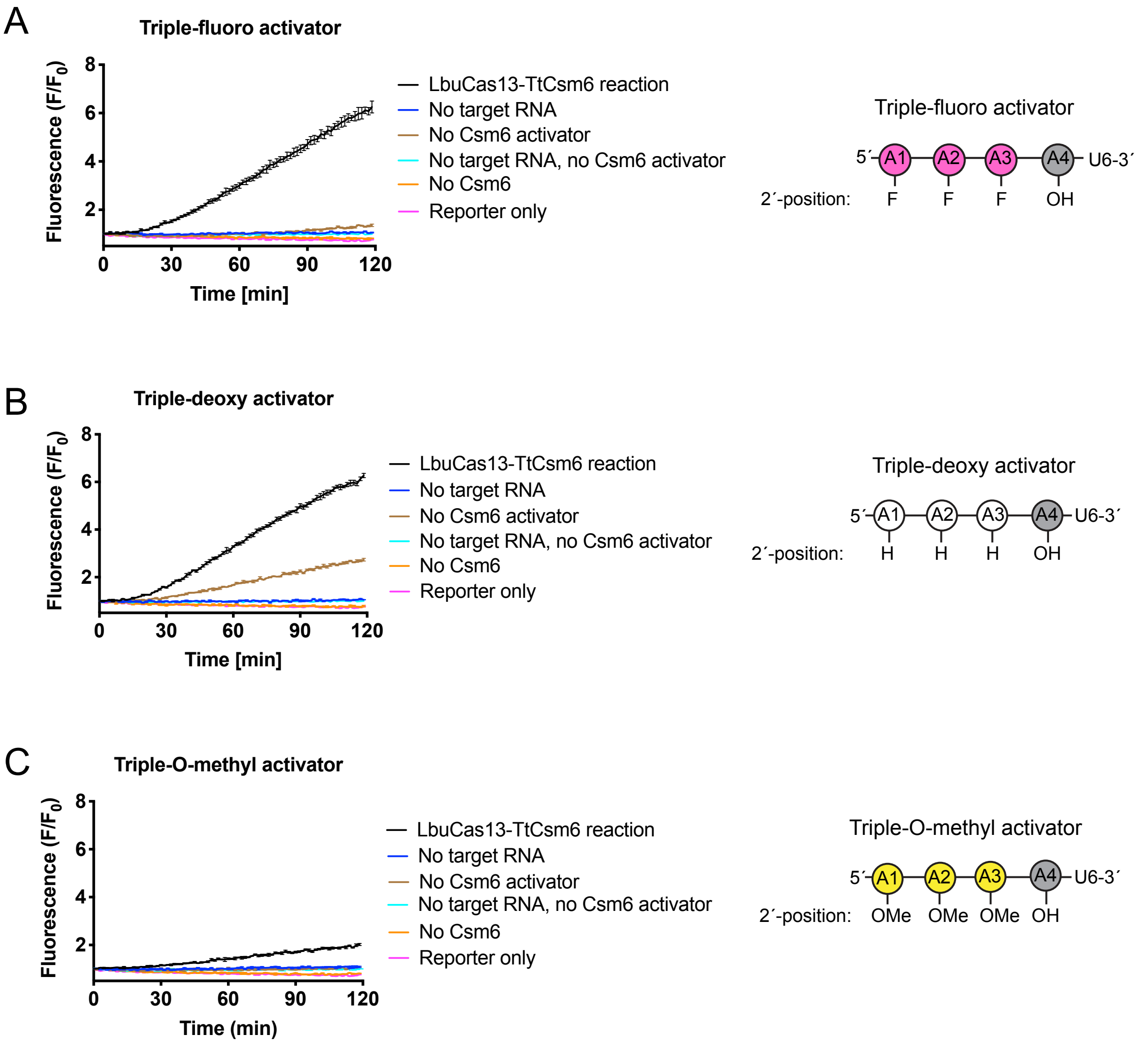

### Extended Data Figure 4

A

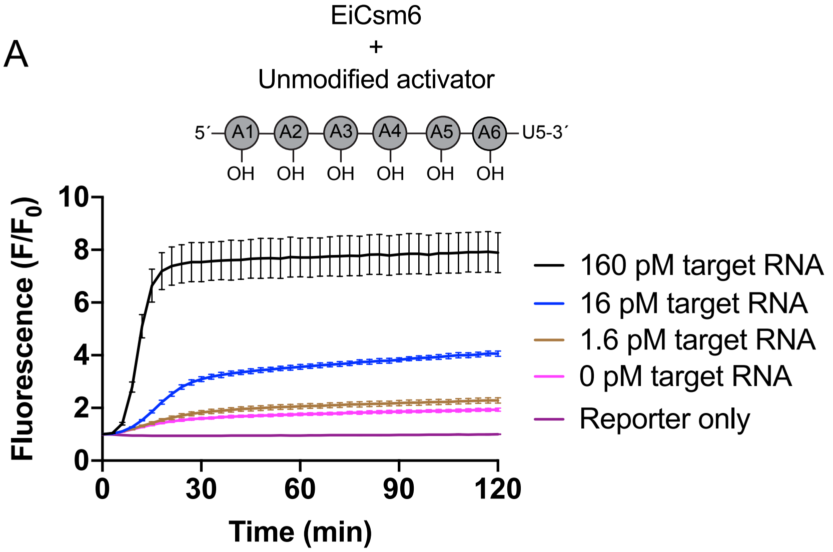


B

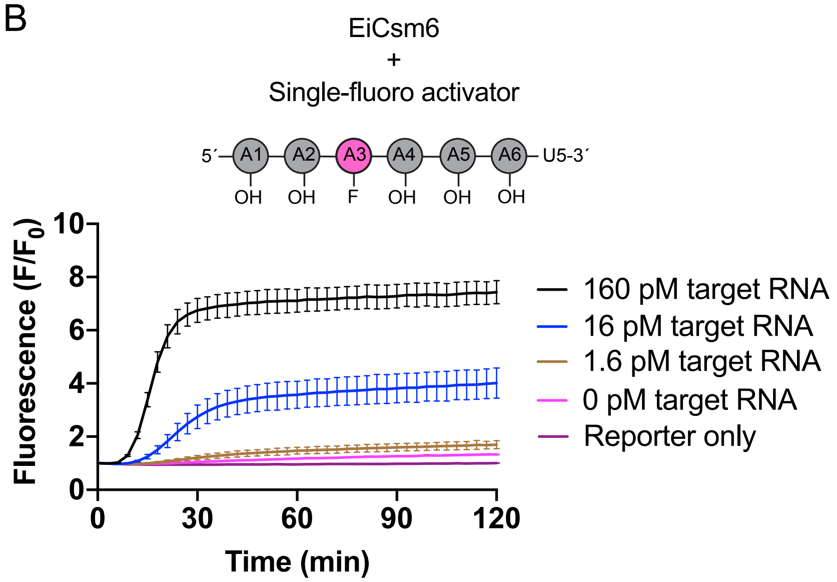

Extended Data Figure 5

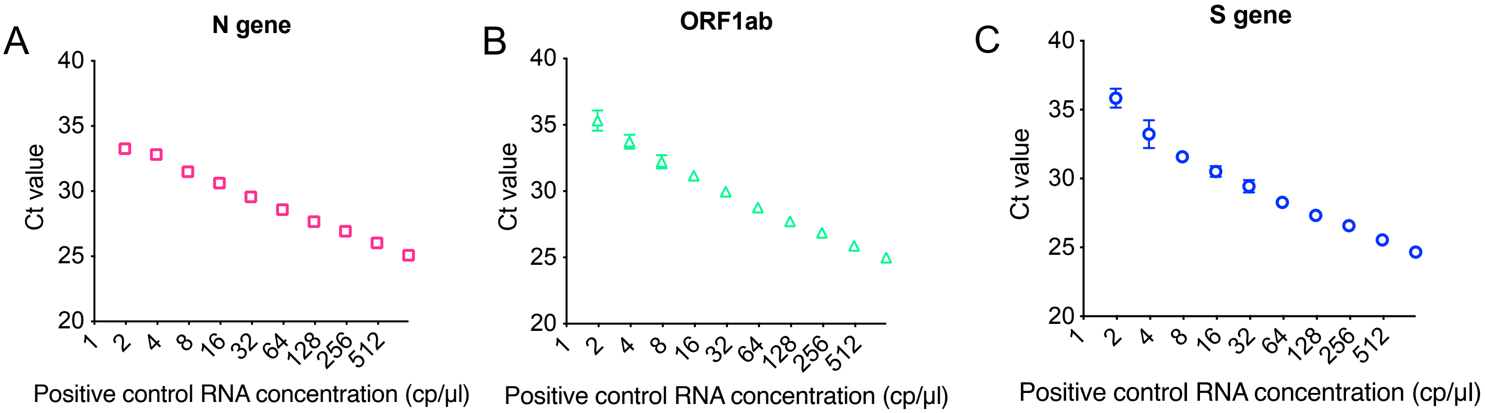

Extended Data Figure 6

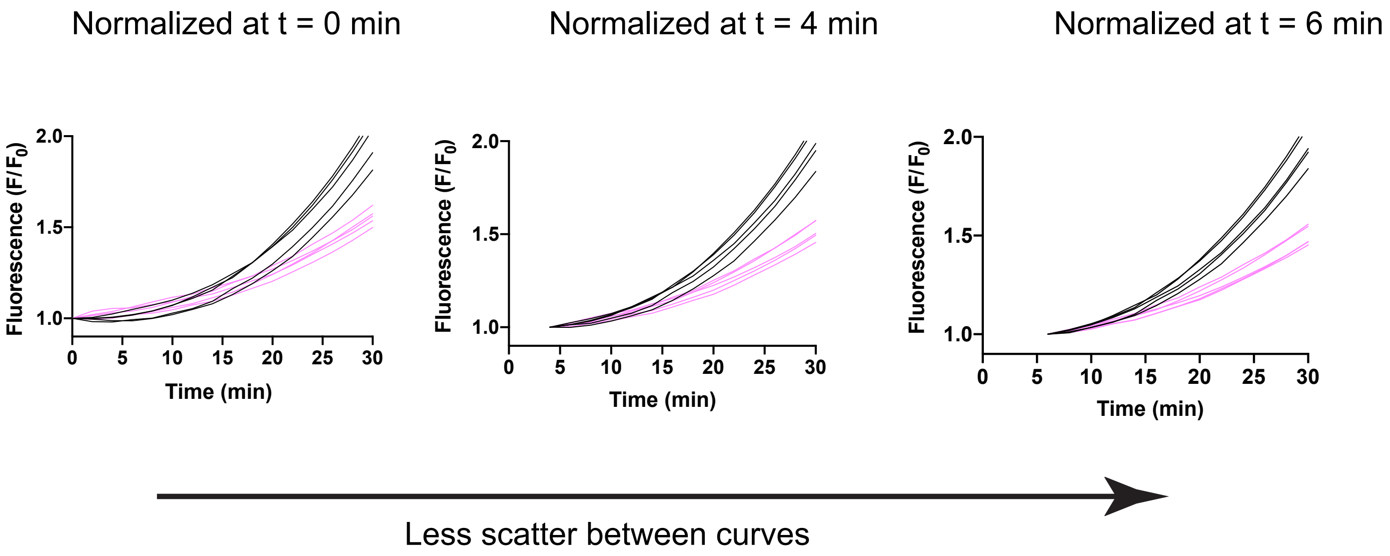
